# Estimating the epidemic reproduction number from temporally aggregated incidence data: a statistical modelling approach and software tool

**DOI:** 10.1101/2022.12.08.22283241

**Authors:** Rebecca K Nash, Anne Cori, Pierre Nouvellet

**Affiliations:** MRC Centre for Global Infectious Disease Analysis, Jameel Institute, School of Public Health, Imperial College London; School of Life Sciences, University of Sussex

## Abstract

**Background:** The time-varying reproduction number (R_t_) is an important measure of epidemic transmissibility; it can directly inform policy decisions and the optimisation of control measures. EpiEstim is a widely used software tool that uses case incidence and the serial interval (SI, time between symptoms in a case and their infector) to estimate R_t_ in real-time. The incidence and the SI distribution must be provided at the same temporal resolution, which limits the applicability of EpiEstim and other similar methods, e.g. for pathogens with a mean SI shorter than the frequency of incidence reporting.

**Methods:** We use an expectation-maximisation algorithm to reconstruct daily incidence from temporally aggregated data, from which R_t_ can then be estimated using EpiEstim. We assess the validity of our method using an extensive simulation study and apply it to COVID-19 and influenza data. The method is implemented in the opensource R package EpiEstim.

**Findings:** For all datasets, the influence of intra-weekly variability in reported data was mitigated by using aggregated weekly data. R_t_ estimated on weekly sliding windows using incidence reconstructed from weekly data was strongly correlated with estimates from the original daily data. The simulation study revealed that R_t_ was well estimated in all scenarios and regardless of the temporal aggregation of the data. In the presence of weekend effects, R_t_ estimates from reconstructed data were more successful at recovering the true value of R_t_ than those obtained from reported daily data.

**Interpretation:** R_t_ can be successfully recovered from aggregated data, and estimation accuracy can even be improved by smoothing out administrative noise in the reported data.

**Funding:** MRC doctoral training partnership, MRC centre for global infectious disease analysis, the NIHR HPRU in Modelling and Health Economics, and the Academy of Medical Sciences Springboard, funded by the AMS, Wellcome Trust, BEIS, the British Heart Foundation and Diabetes UK.

## Introduction

As infectious disease outbreaks become more common, it is increasingly important to rapidly characterise the threat of emerging and re-emerging pathogens.^1^ Transmissibility, i.e. a pathogen’s ability to spread through a population, can be quantified using the time-varying reproduction number, R_t_, defined as the average number of infections that are caused by a primary case at time t of an outbreak. R_t_ signals whether an outbreak is growing (R_t_ > 1) or declining (R_t_ < 1), and whether current interventions are sufficient to control the spread of the disease.

One of the most popular tools for real-time R_t_ estimation, the R package EpiEstim, relies on observing the incidence data and supplying an estimated serial interval (SI) distribution – the time between symptom onset in a case and their infector. EpiEstim requires that the SI distribution and incidence data are supplied using the same time units. This can be problematic when daily incidence data is not reported, which is common for many diseases, such as influenza, Zika virus disease, and most notifiable diseases in countries such as the UK and the US.^2–5^ Additionally, several studies intentionally aggregate data to reduce the impact of daily reporting variability; administrative noise, such as “weekend effects”, are characterised by a drop in reported cases over weekends, due to reduced care seeking and longer delays in reporting, followed by a peak on Mondays.^6,7^ A commonly used workaround is to aggregate the SI distribution to match the frequency of incidence reporting,^8,9^ however this is not possible if the SI is shorter than the aggregation of data. For example, influenza-like illness is typically reported on a weekly basis, but influenza has an estimated mean SI of 2-4 days.^10,11^ Similarly, reporting of COVID-19, which has an estimated SI of 3-7 days, has typically moved from daily to weekly.^12,13^ Therefore, enabling estimation of R_t_ from temporally aggregated data is critical to ensure methods such as EpiEstim are widely applicable.^14^

In this study, we combine an expectation-maximisation (EM) algorithm with the renewal equation approach implemented in EpiEstim to reconstruct daily incidence from aggregated data and estimate R_t_. We assess the performance of the method using influenza and COVID-19 data, in addition to an extensive simulation study.

## Methods

### EpiEstim

EpiEstim uses the renewal equation (eq.1), a form of branching process model.^15^ In this formulation, the incidence of new symptomatic cases at time t (I_t_) is approximated by a Poisson process, where I_t-s_ is the past incidence, and *g*_s_ is the probability mass function of the serial interval.

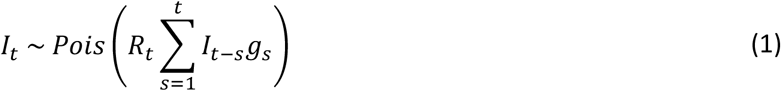

With EpiEstim, R_t_ can be assumed to remain constant within user defined time windows, which smooth out estimates.

### Extending EpiEstim for coarsely aggregated data

We extended EpiEstim to estimate R_t_ from aggregated incidence data (I_w_), where each aggregation window (w) is >1 day, whilst still conditioning on an assumed serial interval distribution (*g*_s_). We use an EM algorithm to iteratively reconstruct daily incidence (I_t_) from I_w_, and in turn estimate R_t_. We present the method with weekly data in mind, but the method and software can be applied to any temporal aggregation (Figure 1 & appendix pp22-24). The algorithm involves three steps: initialisation, expectation, and maximisation.

**Figure 1.**
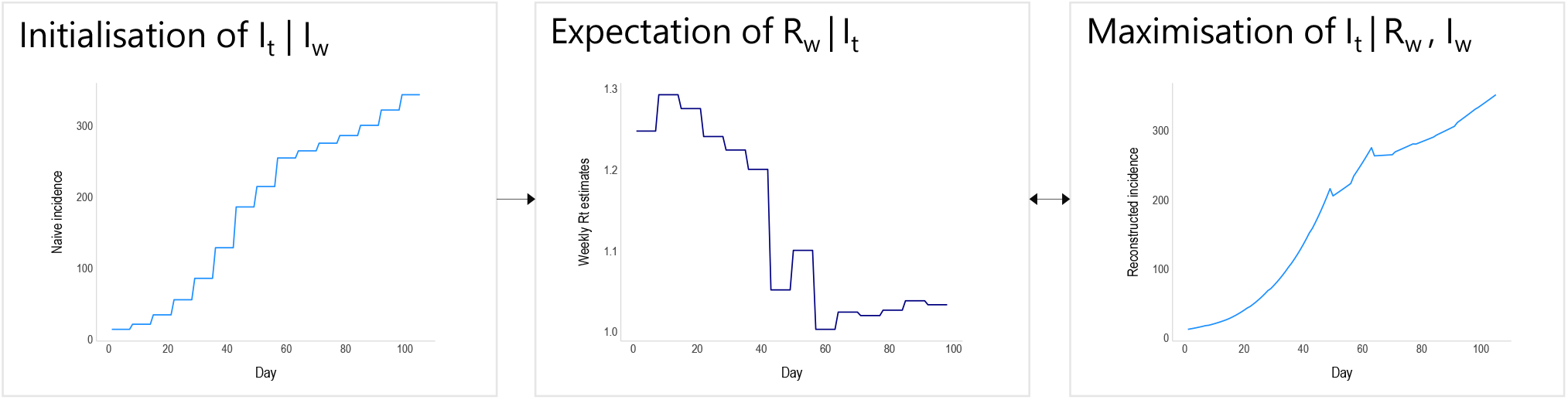
Schematic of the EM algorithm approach used to reconstruct daily incidence (I_t_) from weekly aggregated incidence data (I_w_). The algorithm is initialised with a naive disaggregation of the weekly incidence (assuming constant daily incidence throughout the aggregation window). The resulting daily incidence is then used to estimate the reproduction number for each aggregation window, in this case for each week, R_w_. R_w_ is converted into a growth rate (see eq. 2), which is in turn used to reconstruct daily incidence data, whilst ensuring that if I_t_ were to be reaggregated it would still sum to the original weekly totals. The process cycles between the expectation and maximisation steps until convergence.

### Initialisation

The algorithm is initialised with naively disaggregated incidence data. For weekly data, the total incidence for each week is split evenly over 7 days (allowing for non-integers).

### Expectation

The current reconstructed I_t_ is used to estimate the expected reproduction number for each aggregation window, R_w_, obtained as the posterior mean from EpiEstim.^16^

### Maximisation

Conditional on R_w_, we reconstruct the most likely I_t_. First, R_w_ is translated into a daily growth rate for that week (r_w_), using Wallinga and Lipsitch’s method:^17^

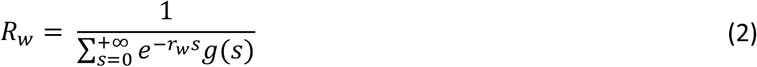

I_t_ for that week is then computed assuming exponential growth, with a multiplying constant k_w_ ensuring that when reaggregated, the reconstructed I_t_ matches the original I_w_:

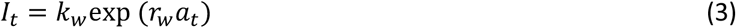

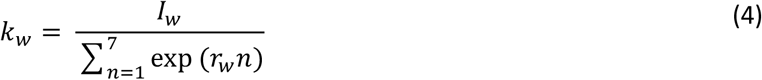

where t is time (in days) and a_t_ is an index representing the day of the aggregation window, e.g. taking values 1 to 7.

The process is repeated iteratively until convergence, at which point I_t_ can be used to estimate the full posterior distribution of R_t_ using EpiEstim. For this final step, R_t_ can be estimated on any time window.

### Case studies

We chose datasets where incidence data was available daily, and then artificially aggregated them to weekly counts. R_t_ was estimated from daily incidence that was reconstructed from weekly aggregated data using our new approach, and compared to R_t_ estimates obtained from the reported daily incidence using the original EpiEstim R package. All R_t_ estimates were made using both daily and weekly sliding time windows, and we refer to those estimates as daily R_t_ estimates and weekly R_t_ estimates respectively.

We considered three characteristics: 1) mean R_t_ estimates, 2) uncertainty in the R_t_ estimates, and 3) the classification of R_t_ as increasing, uncertain or declining (appendix pp8-9). To compare the performance of this approach to the original method, we assessed the correlations between each of the three characteristics when using the reported and reconstructed incidence.

The priors for R_w_ and R_t_ were set to a mean and standard deviation of 5.

### Influenza

We obtained a five-week subset of a dataset (11^th^ December 2009 – 14^th^ January 2010) on US active component military personnel (employed by the military as their full-time occupation) that made an outpatient visit to a permanent military treatment facility describing a respiratory-related illness. This daily incidence by date of presentation at a clinic was originally obtained by Riley et al. from the Armed Forces Health Surveillance Center and were digitally extracted for use here.^18^ We used a mean SI of 3.6 days and SD of 1.6 days.^10^

### COVID-19

Incidence of UK COVID-19 cases and deaths were taken from the UK government website.^19^ For COVID-19 cases, we obtained ninety-seven weeks of data (21^st^ February 2020 to 30^th^ December 2021) for incidence by date of specimen, which is the date that a sample was taken from an individual which later tested positive. For COVID-19 deaths, we used ninety-six weeks of data (2^nd^ March 2020 to 2^nd^ January 2022) for incidence by date of death within twenty-eight days of a positive test. We assumed a mean SI of 6.1 days and SD of 4.2 days.^12^

### Simulation study

We considered scenarios where R_t_ either remained constant or varied over time, with a stepwise or gradual change. For each scenario, one hundred seventy-day epidemic trajectories were simulated using a Poisson branching process as implemented in the R package projections.^20^ Daily datasets were aggregated weekly and used to estimate R_t_ using the proposed method; these values were compared to R_t_ estimates obtained from simulated daily data using the original EpiEstim R package. We explored the impact of weekend effects on R_t_ estimates, the ability to supply alternative temporal aggregations of data e.g., three-day, ten-day, or two-weekly aggregations, and finally, the number of iterations required to reach convergence when reconstructing daily incidence data. The full simulation study description and details can be found in the appendix.

## Results

Hereafter, we refer to reported and reconstructed incidence data, these are the reported daily incidence and the daily incidence that has been reconstructed from weekly aggregated data, respectively.

### Influenza

The reconstructed incidence of influenza was much smoother than the reported incidence, which showed clear weekend effects and lower reported cases on two public holidays, both occurring on Fridays (Figure 2A & appendix p8). Considering weekly sliding R_t_ first, there was a high correlation in both the mean R_t_ estimates derived from each dataset (R^2^ = 0.91, Figure 2C & appendix p2) and their associated uncertainty (R^2^ = 0.93 & Figure 2C). The overall agreement in the classification of R_t_ reached 81.8% (see methods & appendix p9).

**Figure 2.**
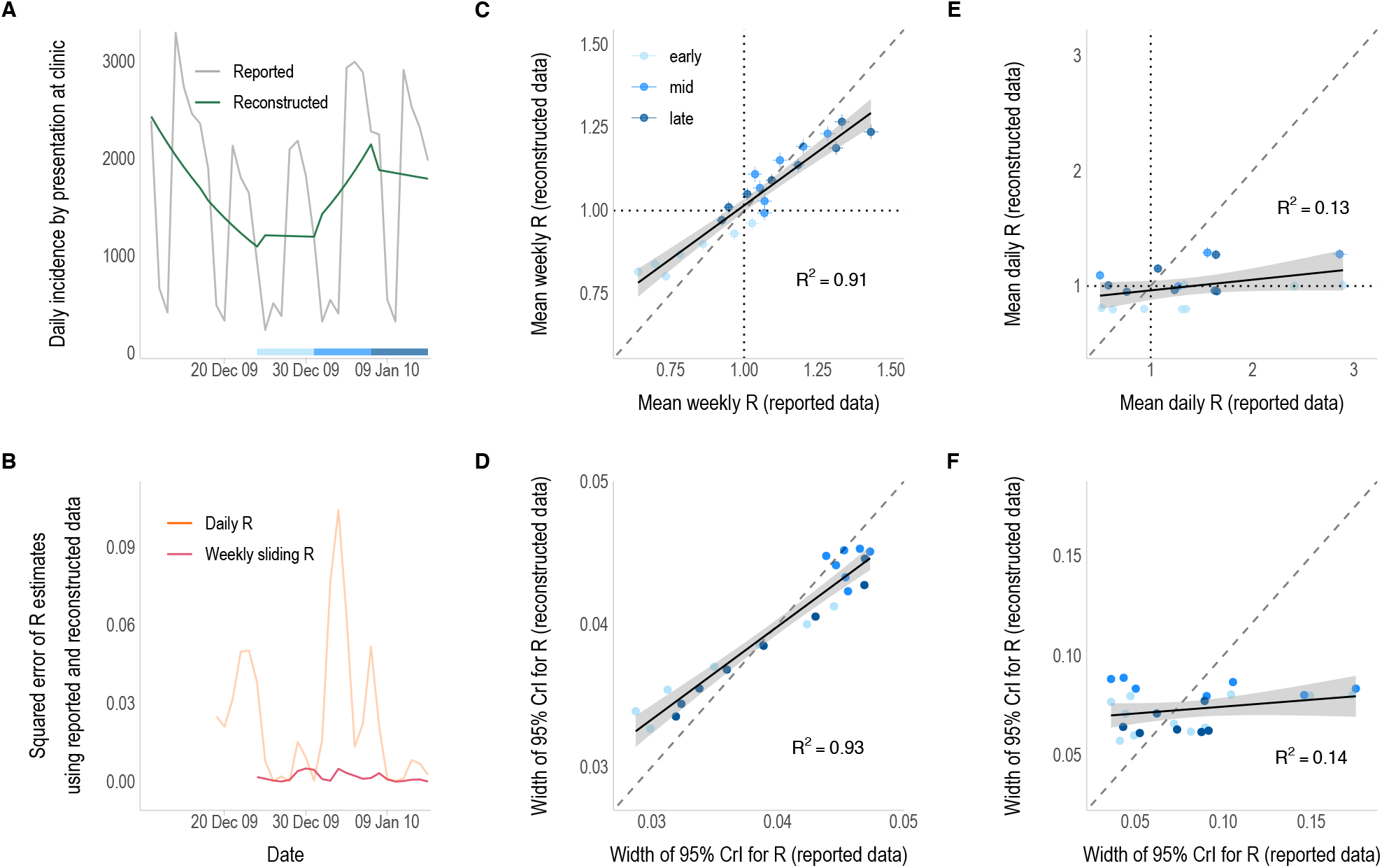
R_t_ estimates from daily incidence that was either reported or reconstructed from weekly aggregated influenza data. A) The reported (grey) and reconstructed (green) daily incidence of influenza by date of presentation at a military clinic. B) Squared error of the daily (orange) and weekly sliding (pink) R_t_ estimates that were made from reconstructed daily data compared to those obtained from the reported daily data. R_t_ estimation starts on the first day of the second aggregation window (day 8 – 18^th^ December 2009) and is plotted on the last day of the time window used for estimation (i.e., starting on day 9 (19^th^ December) for daily estimates and day 14 (24^th^ December) for weekly estimates). Note: the x-axis is shared with the incidence plot above. C & E) Correlation between the weekly sliding (C) and daily (E) mean R_t_ estimates using reconstructed data (y-axis) and reported daily data (x-axis). Vertical and horizontal lines depict the 95% credible intervals and dotted lines show the threshold of R_t_ = 1. D & F) Correlation between the uncertainty in the weekly sliding (D) and daily (F) R_t_ estimates, defined as the width of the 95% credible intervals, using the reconstructed (y-axis) and reported (x-axis) daily data. The colour of the points in panels C-F correspond to the epidemic phase, i.e. the early (19^th^ – 30^th^ December for daily estimates, or 24^th^ – 30^th^ December for weekly sliding estimates), middle (31^st^ December – 6^th^ January) or late (7^th^ – 14^th^ January) phase of the data, shown by the strip in panel A. Solid lines show the linear model fit with 95% confidence intervals (grey shading). Dashed lines represent the x = y line.

In contrast, mean daily R_t_ estimates differed markedly depending on whether the reported or reconstructed data were used, with an R^2^ of 0.13 and much higher mean R_t_ and uncertainty in estimates obtained from reported data (Figure 2E-F). Higher mean R_t_ estimates coincided with large peaks in the reported daily incidence (typically on Mondays), as daily R_t_ estimates were not smoothed and therefore more affected by intra-weekly variability (appendix p2). The overall agreement in the classification of daily R_t_ estimates was much lower, with only 44.4% agreement (appendix p9).

In this case study, the greatest differences in R_t_ estimates tended to correspond to time periods when the reported and reconstructed incidence data were most dissimilar (Figure 2B & appendix p3). There was no apparent pattern in the estimates with regard to the outbreak phase, i.e. early, mid or late-phase, but this is likely due to this dataset being a snapshot of incidence taken from within an established epidemic (Figure 2).

### COVID-19 cases

The reconstructed incidence of COVID-19 smoothed out intra-weekly variability, caused by factors such as weekend effects (Figure 3A & appendix pp7-8). Weekly sliding R_t_ estimates obtained from reconstructed and reported incidence were similar, both in their means (R^2^ = 0.98) and their level of uncertainty (R^2^ = 0.99, Figure 3C-D & appendix p4). Mean daily R_t_ estimates were less well correlated (R^2^ = 0.67), although the difference is less marked than in the influenza case study (Figure 3E), and the uncertainty in the estimates was similar across both approaches (R^2^ = 0.97, Figure 3F). Most of the discrepant R_t_ estimates and higher levels of uncertainty coincide with the early phase of the outbreak when incidence was lower (Figure 3E-F). Outside of periods of low incidence, the largest differences in R_t_ estimates tended to correspond to time periods with greater disparities between the reported and reconstructed incidence data (Figure 3B & appendix p5). The overall agreement in the classification of R_t_ estimates was higher than for influenza, with 74.4% and 94.9% agreement for daily and weekly sliding R_t_ estimates respectively (appendix p9).

**Figure 3.**
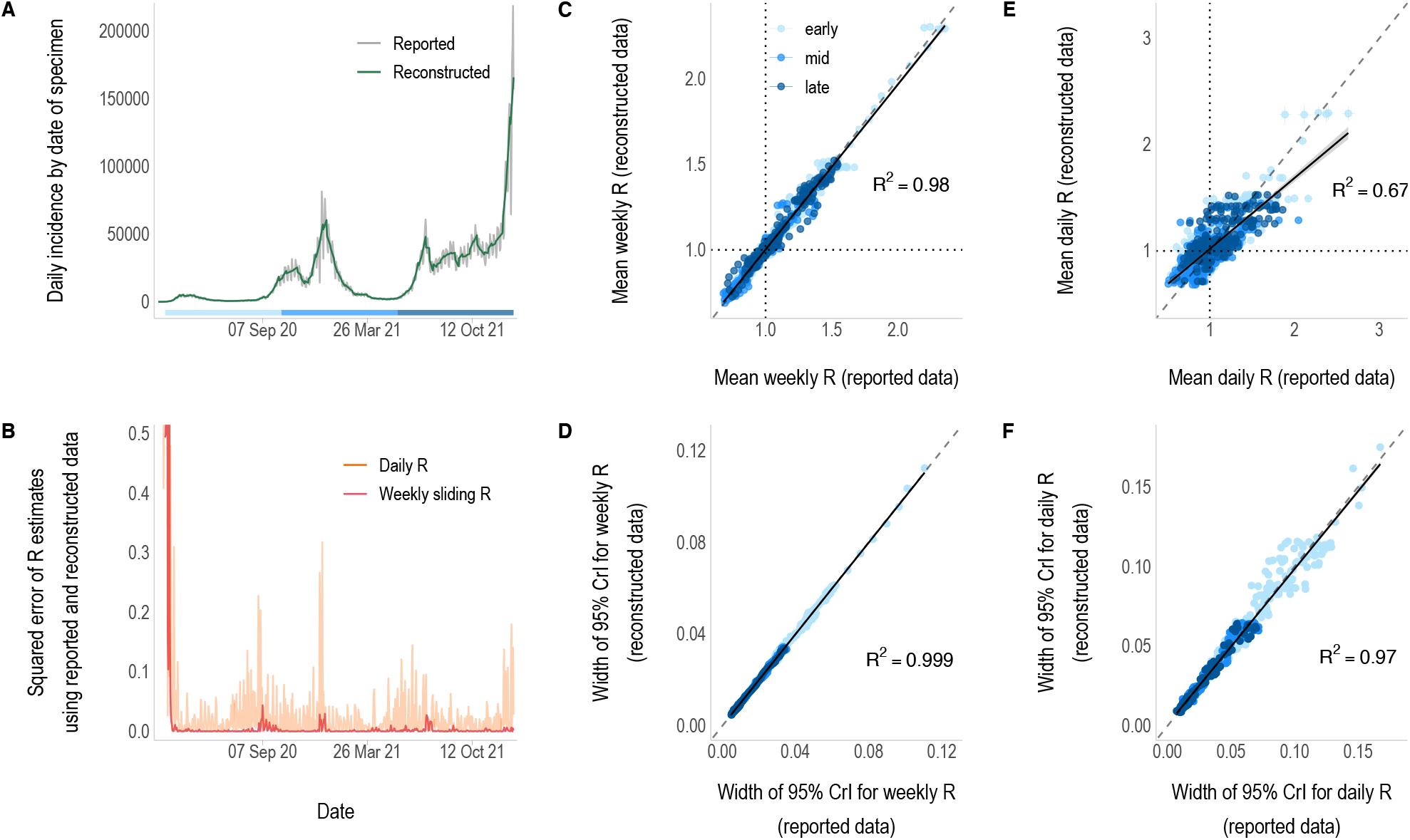
R_t_ estimates from daily incidence that was either reported or reconstructed from weekly aggregated COVID-19 case data. A) The reported (grey) and reconstructed (green) daily incidence of COVID-19 by date of specimen. B) Squared error of the daily (orange) and weekly sliding (pink) R_t_ estimates made from reconstructed data compared to those obtained from the reported daily data. R_t_ estimation starts on the first day of the second aggregation window (day 8 – 28^th^ February 2020) and is plotted on the last day of the time window used for estimation (i.e., starting on day 9 (29^th^ February) for daily estimates and day 14 (5^th^ March) for weekly estimates). Note: the x-axis is shared with the incidence plot above and the y-axis has been limited to 0.5 for clarity. C & E) Correlation between the weekly sliding (C) and daily (E) mean R_t_ estimates using reconstructed (y-axis) and reported (x-axis) daily data, excluding the first 30 days due to low incidence. Vertical and horizontal lines depict the 95% credible intervals and dotted lines show the threshold of R_t_ = 1. D & F) Correlation between the uncertainty in the weekly sliding (D) and daily (F) R_t_ estimates, defined as the width of the 95% credible intervals, using the reconstructed (y-axis) and reported (x-axis) daily data. The colour of the points in panels C-F correspond to the epidemic phase, i.e. the early (21^st^ March – 12^th^ October 2020), middle (13^th^ October 2020 – 22^nd^ May 2021) or late (23^rd^ May – 30^th^ December 2021) phase of the data, shown by the strip in panel A. Solid lines show the linear model fit with 95% confidence intervals (grey shading). Dashed lines represent the x = y line.

### COVID-19 deaths

The reported incidence of COVID-19 deaths was much less influenced by day-to-day variation. The reconstructed daily incidence was more similar to the observed daily data than in the previous case studies (Figure 4A). Both weekly and daily R_t_ estimates obtained from weekly data were highly consistent with those obtained from daily observations (R^2^ = 0.98 and R^2^ = 0.80 respectively, Figure 4C & 4E). The overall agreement in R_t_ classifications for daily estimates was the highest of all case studies at 85.8%, and 93.3% for weekly R_t_ estimates (appendix p9). Discrepancies between the two mostly coincide with periods of particularly low incidence of deaths (Figure 4B & appendix p7). The overall lower incidence of COVID-19 deaths compared to COVID-19 cases means there is greater uncertainty in R_t_ estimates in this case study (Figure 4D, 4F & appendix p6). However, there was minimal difference in the uncertainty of estimates obtained from daily and weekly data (Figure 4D & 4F).

**Figure 4.**
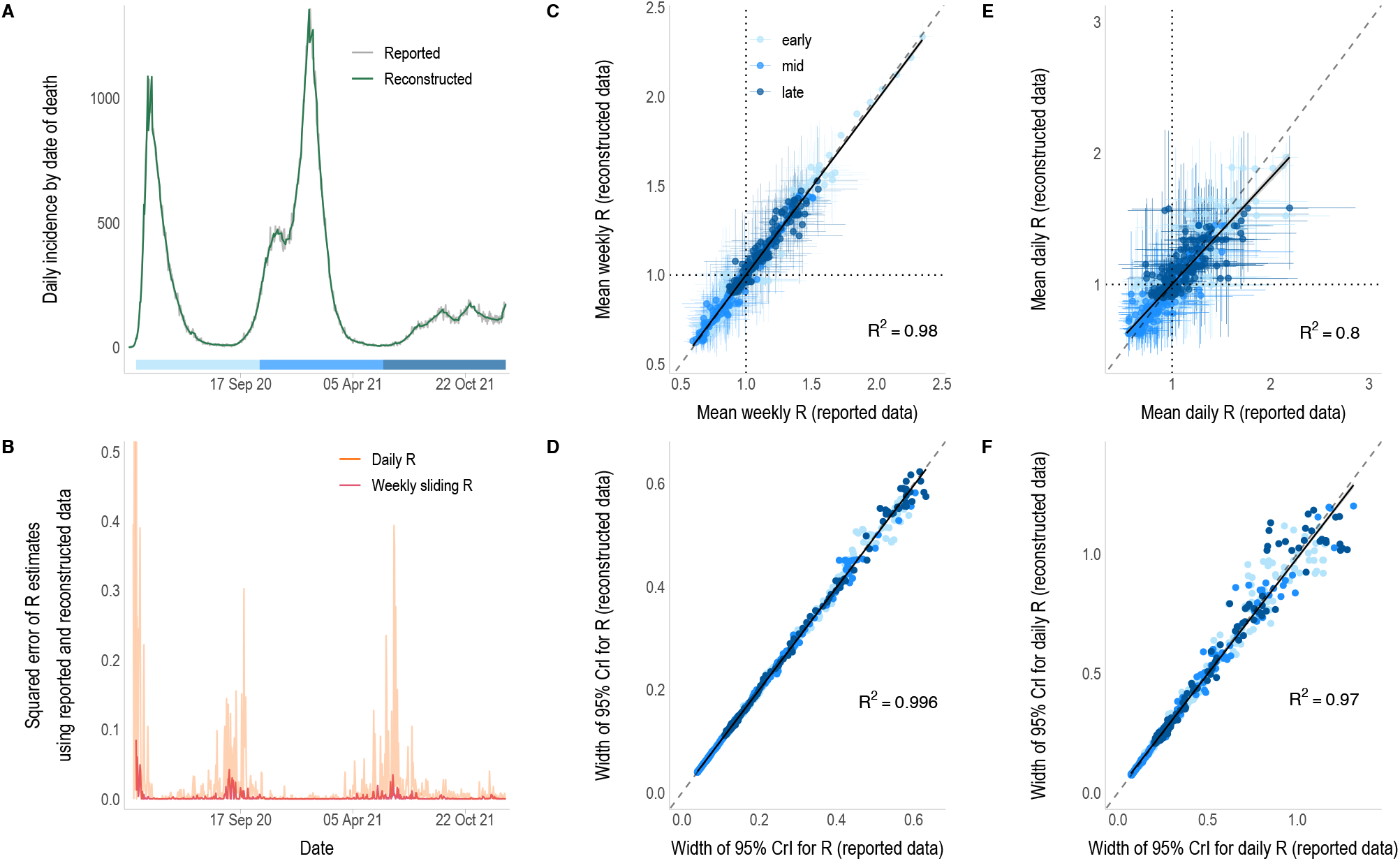
R_t_ estimates from daily incidence that was either reported or reconstructed from weekly aggregated COVID-19 death data. A) The reported (grey) and reconstructed (green) daily incidence of COVID-19 by date of death within 28 days of a positive test. B) Squared error of the daily (orange) and weekly sliding (pink) R_t_ estimates that were made from reconstructed data compared to those obtained from the reported daily data. R_t_ estimation starts on the first day of the second aggregation window (day 8 – 9^th^ March 2020) and is plotted on the last day of the time window used for estimation (i.e., starting on day 9 (10^th^ March) for daily estimates and day 14 (15^th^ March) for weekly estimates). Note: the x-axis is shared with the incidence plot above and the y-axis has been limited to 0.5 for clarity. C & E) Correlation between the weekly sliding (C) and daily (E) mean R_t_ estimates using reconstructed (y-axis) and reported daily data (x-axis), excluding the first 30 days due to low incidence. Vertical and horizontal lines depict the 95% credible intervals and dotted lines show the threshold of R_t_ = 1. D & F) Correlation between the uncertainty in the weekly sliding (D) and daily (F) R_t_ estimates, defined as the width of the 95% credible intervals, using the reconstructed (y-axis) and reported daily (x-axis) data. The colour of the points in panels C-F correspond to the epidemic phase, i.e. the early (31^st^ March - 20^th^ October 2020), middle (21^st^ October 2020 – 28^th^ May 2021) or late (29^th^ May 2021 – 2^nd^ January 2022) phase of the data, shown by the strip in panel A. Solid lines show the linear model fit with 95% confidence intervals (grey shading). Dashed lines represent the x = y line.

In all case-studies, incidence reconstructions converged within 10 iterations of the EM algorithm. The overall process of R_t_ estimation from weekly aggregated data took three seconds or less to run on MacOS (2 GHz Quad-Core Intel Core i5) 16GB RAM (appendix p10); the influenza scenario, with over 57,000 cases, took two seconds to run, whilst the COVID-19 cases and deaths scenarios, with an overall incidence over 149,000 and 13 million cases respectively, took three seconds to run.

### Simulation study

The method performed well across all scenarios, successfully estimating R_t_ from the aggregated simulated data (appendix pp10-20). Convergence of the EM algorithm was quick, with negligible differences in the reconstructed incidence beyond 5 iterations (appendix p22).

When introducing weekend effects into simulated data, R_t_ estimates from reconstructed incidence were more successful at recovering the true value of R_t_ than when using reported incidence (appendix p21). The method can also be successfully applied to other temporal aggregations of data, e.g. three-, ten- or fourteen-day windows (appendix pp22-24).

## Discussion

Estimates of the time-varying reproduction number (R_t_) have frequently been used to inform and guide policymaking during outbreaks, and a commonly used approach to estimate R_t_ is EpiEstim, which relies on daily incidence data. However, maintaining daily incidence databases requires substantial time and investment in resources, which is not always feasible, particularly for less acute or routinely reported diseases. Therefore, in practice, many diseases are not reported on a daily basis, including influenza and other notifiable diseases in the UK and US.^2–5^ As the COVID-19 pandemic persists, daily reporting is also becoming less common.^21^ Coarsely aggregated data can be challenging to deal with in the context of R_t_ estimation methods, restricting their applications in certain contexts. In this study, we develop a statistical framework and tool that allows R_t_ estimation from aggregated incidence without introducing bias. Using influenza and COVID-19 data, alongside a simulation study, we demonstrate how a simple expectation-maximisation algorithm approach can rapidly reconstruct daily incidence data and accurately estimate R_t_.

In all case studies, direct comparisons between weekly sliding R_t_ estimates show that very similar estimates can be made from the reported daily incidence and the reconstructed daily incidence from weekly aggregated data. However, daily R_t_ estimates are more influenced by noise, such as intra-weekly variability, leading to greater disparities in estimates between datasets. There are clear weekend effects exhibited in the influenza and COVID-19 case data (appendix p8), leading to peaks and troughs in the reported incidence and the resulting daily R_t_ estimates (Figures 2 & 3, appendix pp2&4). Using reconstructed incidence considerably smoothed the daily R_t_ estimates, removing the impact of weekend-effects. The overall agreement in the classification of R_t_ as increasing, uncertain, or declining between estimates made from each dataset rose substantially when some of the variability in the reported data was smoothed by estimating R_t_ using weekly sliding windows (appendix pp8-9).

Despite both being affected by weekly periodicity in reporting, concordance of R_t_ estimates obtained from COVID-19 case data is considerably better than for influenza, perhaps due to the greater quantity of data, with a very strong positive correlation between daily and weekly R_t_ estimates (Figure 3). This is reflected in the high overall agreement in the classification of R_t_ estimates obtained from the reported and reconstructed datasets. It is important to note that outlying and much larger R_t_ estimates obtained from both datasets coincide with the early phase of the epidemic, when incidence was lower and the prior for R_t_ (μ=5, σ=5) had more weight on estimates.

During the early stages of epidemics, despite there being far fewer deaths than cases, death data can sometimes be considered more reliable.^22,23^ For example, case reporting is affected by surveillance system quality and the robustness of testing practices, which can vary considerably over the course of an epidemic, especially early on. COVID-19 incidence by date of death is much less influenced by administrative noise in the data (appendix p8), and the reconstructed incidence is most similar to the reported daily incidence of any case study. Therefore, the greatest differences in R_t_ estimates from death data coincide with periods of low incidence (appendix p7) when uncertainty increases. Weekly sliding R_t_ estimates are equally as correlated as those from COVID-19 case data, but daily R_t_ estimates are the most strongly correlated of any dataset (Figure 4). Additionally, there is very high overall agreement in the classification of daily and weekly R_t_ (appendix p9). This provides further support that differences between daily R_t_ estimates for influenza and COVID-19 cases is likely due to the reconstructed incidence smoothing out weekly periodicity in reporting.

To investigate further, weekend effects were artificially introduced to data in the simulation study (appendix p21). We have shown that, when using reported incidence, R_t_ estimates are all strongly influenced by weekend effects (regardless of the smoothing time-window). Reconstructing daily incidence from weekly data completely removes the effect of noise from resulting R_t_ values, greatly improving the accuracy of estimates. This demonstrates that it may be beneficial to artificially aggregate daily data, as has been done in previous studies.^6,7^ However, we did assume quite an extreme level of administrative noise, so in instances where the pattern is less prominent, it may have less of an impact on estimates. Disentangling important temporal trends in R_t_ from noise in the data can be difficult, and if aggregated data is used it will be at the cost of reduced temporal resolution in R_t_ estimates.

This can be seen when the method is applied to data aggregated over longer timescales, such as ten- to fourteen-days (appendix pp22-24). This approach requires two layers of smoothing: 1) the incidence is smoothed over each aggregation window during the reconstruction process and 2) R_t_ estimates are smoothed by the sliding window chosen by the user. If a change in R_t_ occurs at the end of an aggregation window (i.e. on the last day), such as a sudden decrease in R_t_ due to a strict lockdown, that change is detected with a lag, corresponding to the length of the sliding window used for R_t_ estimation (appendix p23). However, if the event occurs mid-aggregation window, then in addition to the usual lag caused by the sliding window, estimates will be affected by the smoothing of the incidence within the aggregation window during reconstruction (appendix p24). The change in R_t_ will seem more gradual over the period that data are aggregated over and will appear to start earlier than in reality (corresponding to the first day of the aggregation window). It is important for users to keep this in mind, particularly when using longer aggregations of data.

Another consideration is that the reconstructed incidence can have discontinuities in the borders between aggregation windows (appendix pp11-12). This occurs because in reconstructing daily incidence we impose that, if it were to be re-aggregated, it would match the original data. Methods that simply fit smoothing splines to weekly data, inferring daily case counts from the daily difference in cumulative counts, are not affected by this.^24,25^ To circumvent this problem, we recommend that sliding windows used to estimate R_t_ are at least equal to or longer than the length of aggregation windows to reduce the impact of discontinuities on estimates (appendix pp22-24).

Alternative approaches include modelling frameworks implemented in the Epidemia and EpiNow2 R packages.^6,22,26^ Daily infections are modelled as a latent process, back-calculated from observed data on cases or deaths, depending on an appropriate infection to observation distribution. In addition, Epidemia integrates further information, such as the infection ascertainment rate (for cases) or the infection fatality rate (for deaths).^22^ This facilitates a ‘nowcasting’ approach, allowing users to estimate R_t_ directly from the unobserved infections, but they typically require more data (e.g. incidence of deaths and cases), more assumptions (e.g. delay distributions and ascertainment rates), and are much more computationally intensive, which can be a barrier to the adoption of such methods by users.^14^

Here, R_t_ estimates are based on a single daily incidence reconstruction, meaning R_t_ can be estimated very rapidly from aggregated data, which is particularly desirable during real-time outbreak analysis.^14^ A potential downside is that uncertainty in R_t_ estimates could be underestimated. However, the simulation study showed that the 95% credible interval of estimates encompassed the correct value of R_t_ the majority of the time, and we found no substantial indication that this approach detrimentally affected our characterisation of the uncertainty.

Given that this method is directly derived from EpiEstim, it relies on similar assumptions and caveats.^15,27^ As time of infection is more difficult to observe than symptom onset, the SI is typically used as an approximation of the generation time in the renewal equation, which may introduce bias.^28^ The SI, the level of undetected cases, and the reporting rate are assumed to remain constant, which is often not the case in practice. Factors such as changes in population immunity, and the introduction of interventions, can alter the SI throughout an epidemic.^29^ Whilst changing case definitions, new testing practices, and increased healthcare-seeking behaviour, can all affect case ascertainment.^15^ Parameters chosen by users can also influence estimation accuracy, for instance, the time window length for temporal smoothing and the prior for R_t_.^27^

To make the method simple to implement for current and future users of EpiEstim, this extension has been fully integrated with the ‘estimate_R()’ function in the original R package on github.^30^ Just one additional parameter is required – the number of days data are aggregated over (with some other optional parameters). More details regarding the applications of this method can be found in the package vignette and associated examples.^30^

## Conclusion

We extended the widely used R_t_ estimation approach proposed by Cori et al.,^15^ and implemented in the R package EpiEstim, to incorporate a new feature which allows R_t_ to be easily estimated from any temporal aggregation of incidence data. We have demonstrated that the method performs well using both simulated and real-world data, recovering or even improving upon the estimates that would have been made from reported daily data. This extension is easy to use and computationally efficient, which will enable epidemiologists and other public health professionals to apply EpiEstim to a wider range of diseases and epidemic contexts.

## Data Availability

The code to apply the method developed is available in the open-source R package EpiEstim: https://github.com/mrc-ide/EpiEstim.

## References

1. Baker RE, Mahmud AS, Miller IF, Rajeev M, Rasambainarivo F, Rice BL, et al. Infectious disease in an era of global change. Nat Rev Microbiol. 2022 Apr;20(4):193–205.

2. National flu and COVID-19 surveillance reports: 2021 to 2022 season [Internet]. GOV.UK. [cited 2022 Jun 27]. Available from: https://www.gov.uk/government/statistics/national-flu-and-covid-19-surveillance-reports-2021-to-2022-season

3. Pacheco O, Beltrán M, Nelson CA, Valencia D, Tolosa N, Farr SL, et al. Zika Virus Disease in Colombia — Preliminary Report. New England Journal of Medicine. 2020 Aug 6;383(6):e44.

4. Notifiable diseases: weekly reports for 2022 [Internet]. GOV.UK. [cited 2022 Jun 27]. Available from: https://www.gov.uk/government/publications/notifiable-diseases-weekly-reports-for-2022

5. Notifiable Infectious Disease Tables | CDC [Internet]. 2021 [cited 2022 Jul 2]. Available from: https://www.cdc.gov/nndss/data-statistics/infectious-tables/index.html

6. Mishra S, Scott J, Zhu H, Ferguson NM, Bhatt S, Flaxman S, et al. A COVID-19 Model for Local Authorities of the United Kingdom [Internet]. medRxiv; 2020 [cited 2022 Jul 1]. p. 2020.11.24.20236661. Available from: https://www.medrxiv.org/content/10.1101/2020.11.24.20236661v1

7. Role of Data Aggregation in Biosurveillance Detection Strategies with Applications from ESSENCE [Internet]. [cited 2022 Jul 2]. Available from: https://www.cdc.gov/mmwr/preview/mmwrhtml/su5301a16.htm

8. Ferguson NM, Cucunubá ZM, Dorigatti I, Nedjati-Gilani GL, Donnelly CA, Basáñez Mg, et al. Countering the zika epidemic in latin america. Science. 2016;353(6297):353–4.

9. Charniga K,Cucunubá ZM, Mercado M, Prieto F, Ospina M, Nouvellet P, et al. Spatial and temporal invasion dynamics of the 2014–2017 Zika and chikungunya epidemics in Colombia. PLOS Computational Biology. 2021 Jul 2;17(7):e1009174.

10. Cowling BJ, Fang VJ, Riley S, Peiris JSM, Leung GM. Estimation of the serial interval of influenza. Epidemiology. 2009 May;20(3):344–7.

11. White LF, Wallinga J, Finelli L, Reed C, Riley S, Lipsitch M, et al. Estimation of the reproductive number and the serial interval in early phase of the 2009 influenza A/H1N1 pandemic in the USA. Influenza and Other Respiratory Viruses. 2009;3(6):267–76.

12. Bi Q, Wu Y, Mei S, Ye C, Zou X, Zhang Z, et al. Epidemiology and transmission of COVID-19 in 391 cases and 1286 of their close contacts in Shenzhen, China: a retrospective cohort study. The Lancet Infectious Diseases. 2020 Aug 1;20(8):911–9.

13. Rai B, Shukla A, Dwivedi LK. Estimates of serial interval for COVID-19: A systematic review and meta-analysis. Clin Epidemiol Glob Health. 2021;9:157–61.

14. Nash RK, Nouvellet P, Cori A. Real-time estimation of the epidemic reproduction number: Scoping review of the applications and challenges. PLOS Digital Health. 2022 Jun 27;1(6):e0000052.

15. Cori A, Ferguson NM, Fraser C, Cauchemez S. A New Framework and Software to Estimate Time-Varying Reproduction Numbers During Epidemics. Am J Epidemiol. 2013 Nov 1;178(9):1505–12.

16. Cori [aut A cre, Cauchemez S, Ferguson NM, Fraser C, Dahlqwist E, et al. EpiEstim: Estimate Time Varying Reproduction Numbers from Epidemic Curves [Internet]. 2021 [cited 2022 Jun 9]. Available from: https://CRAN.R-project.org/package=EpiEstim

17. Wallinga J, Lipsitch M. How generation intervals shape the relationship between growth rates and reproductive numbers. Proceedings of the Royal Society B: Biological Sciences. 2007 Feb 22;274(1609):599–604.

18. Riley P, Cost AA, Riley S. Intra-Weekly Variations of Influenza-Like Illness in Military Populations. Military Medicine. 2016 Apr 1;181(4):364–8.

19. Cases in the UK | Coronavirus in the UK [Internet]. [cited 2022 Jan 9]. Available from: https://coronavirus.data.gov.uk/details/cases

20. Jombart T, Nouvellet P, Bhatia S, Kamvar ZN, Taylor T, Ghozzi S. projections: Project Future Case Incidence [Internet]. 2021 [cited 2022 Jun 9]. Available from: https://CRAN.R-project.org/package=projections

21. CSSEGISandData. COVID-19 Data Repository by the Center for Systems Science and Engineering (CSSE) at Johns Hopkins University [Internet]. 2022 [cited 2022 Jul 5]. Available from: https://github.com/CSSEGISandData/COVID-19

22. Flaxman S, Mishra S, Gandy A, Unwin HJT, Mellan TA, Coupland H, et al. Estimating the effects of non-pharmaceutical interventions on COVID-19 in Europe. Nature. 2020;584(7820):257–61.

23. Nouvellet P, Bhatia S, Cori A, Ainslie KEC, Baguelin M, Bhatt S, et al. Reduction in mobility and COVID-19 transmission. Nat Commun. 2021 Feb 17;12(1):1090.

24. Yamauchi T, Takeuchi S, Yamano Y, Kuroda Y, Nakadate T. Estimation of the effective reproduction number of influenza based on weekly reports in Miyazaki Prefecture. Scientific reports. 2019;9(1):1–9.

25. Nishiura H, Chowell G. Early transmission dynamics of Ebola virus disease (EVD), West Africa, March to August 2014. Eurosurveillance. 2014 Sep 11;19(36):20894.

26. Abbott S, Hellewell J, Thompson RN, Sherratt K, Gibbs HP, Bosse NI, et al. Estimating the time-varying reproduction number of SARS-CoV-2 using national and subnational case counts. Wellcome Open Res. 2020 Jun 1;5:112.

27. Gostic KM, McGough L, Baskerville EB, Abbott S, Joshi K, Tedijanto C, et al. Practical considerations for measuring the effective reproductive number, Rt. PLOS Computational Biology. 2020 Dec 10;16(12):e1008409.

28. Britton T, Scalia Tomba G. Estimation in emerging epidemics: biases and remedies. Journal of The Royal Society Interface. 2019 Jan 31;16(150):20180670.

29. Ali ST, Wang L, Lau EHY, Xu XK, D. Z, Wu Y, et al. Serial interval of SARS-CoV-2 was shortened over time by nonpharmaceutical interventions. Science. 2020 Aug 28;369(6507):1106–9.

30. mrc-ide/EpiEstim: A tool to estimate time varying instantaneous reproduction number during epidemics [Internet]. [cited 2022 Aug 9]. Available from: https://github.com/mrc-ide/EpiEstim

