## Supplementary Information for "Estimating the epidemic reproduction number from temporally aggregated incidence data: a statistical modelling approach and software tool"

### 1. Overview

In this supplementary information we present further results from our analysis of influenza-like illness and COVID-19 data. We also present analysis from a simulation study, which was used to validate our method using a variety of simulated epidemic scenarios, where $R_t$ either remained constant or varied over time. As part of the simulation study, we explored the impact of weekend effects on estimates of $R_t$, the ability to supply alternative temporal aggregations of data e.g., 3-day, 10-day, or two-weekly aggregations, and finally, we discuss the number of iterations generally required to reach convergence when reconstructing daily incidence data using our Expectation-Maximisation (EM) algorithm.

### 2. Additional results

As described in the main text, reported incidence data for cases of influenza-like illness, COVID-19 cases, and COVID-19 deaths was used to estimate $R_t$ using the original EpiEstim R package[1] as well as our extended method. The reported daily data were artificially aggregated to a weekly timescale to replicate a typical scenario where data are reported on a weekly basis. Both the reported daily data and the reconstructed daily data, which was obtained from the weekly aggregations of incidence using our EM algorithm, were used to estimate $R_t$ over daily and weekly sliding windows ending on day t.

#### a. Influenza

In the influenza case study, both the daily and weekly sliding $R_t$ estimates are smoother when estimated from the reconstructed data as opposed to the reported data (Figure S1). The prominent weekly oscillations in the daily $R_t$ estimates from the reported data are likely due to the impact of day-to-day variations in reporting, such as weekend effects, which in this dataset may be caused by factors including the military clinic's opening hours or the military personnel's working hours.[2] These intra-weekly fluctuations are lost once the data are aggregated; therefore, the reconstructed incidence and the $R_t$ estimates based on it will be smoother and less affected by this variability (Figure S1 & Figure S2A-B).

Weekly sliding $R_t$ estimates account for some of this variation, leading to smoother $R_t$ estimates from both the reported and reconstructed data (Figure S1B). It is much clearer here that the estimates follow the same general trend, but once again, the $R_t$ estimates are slightly smoother from the reconstructed data.

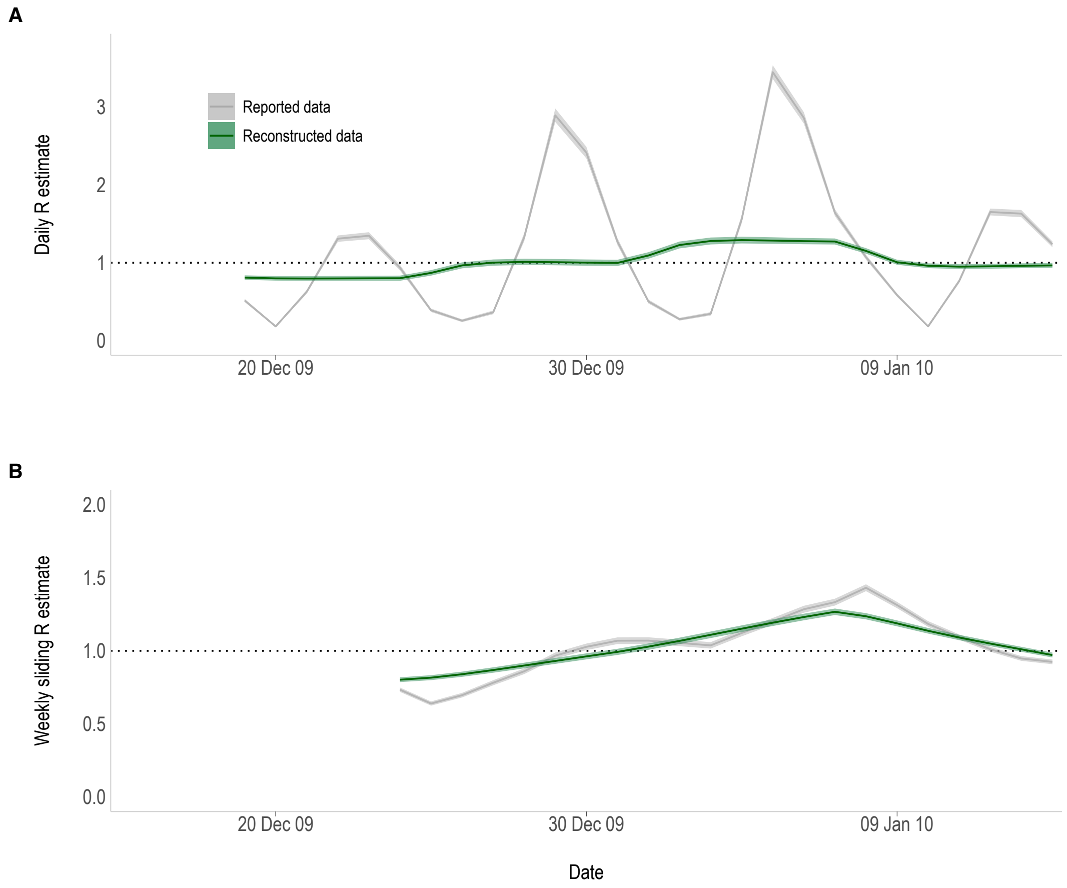

**Figure S1.** The A) daily and B) weekly sliding $R_t$ estimates for influenza based on the reported (grey) and the reconstructed (green) daily data by date of presentation at the military clinic. $R_t$ estimates start on the first day of the second aggregation window (day 8 – 18th December 2009) and are plotted at the end of the time window. Shading corresponds to the 95% credible interval of the estimates.

Bias in the $R_t$ estimates was assessed by computing the absolute difference in the estimates from the reported and reconstructed data (Figure S2C-D). The greater differences between the $R_t$ estimates correspond to the largest disparities between the reported and reconstructed incidence data (Figure S2).

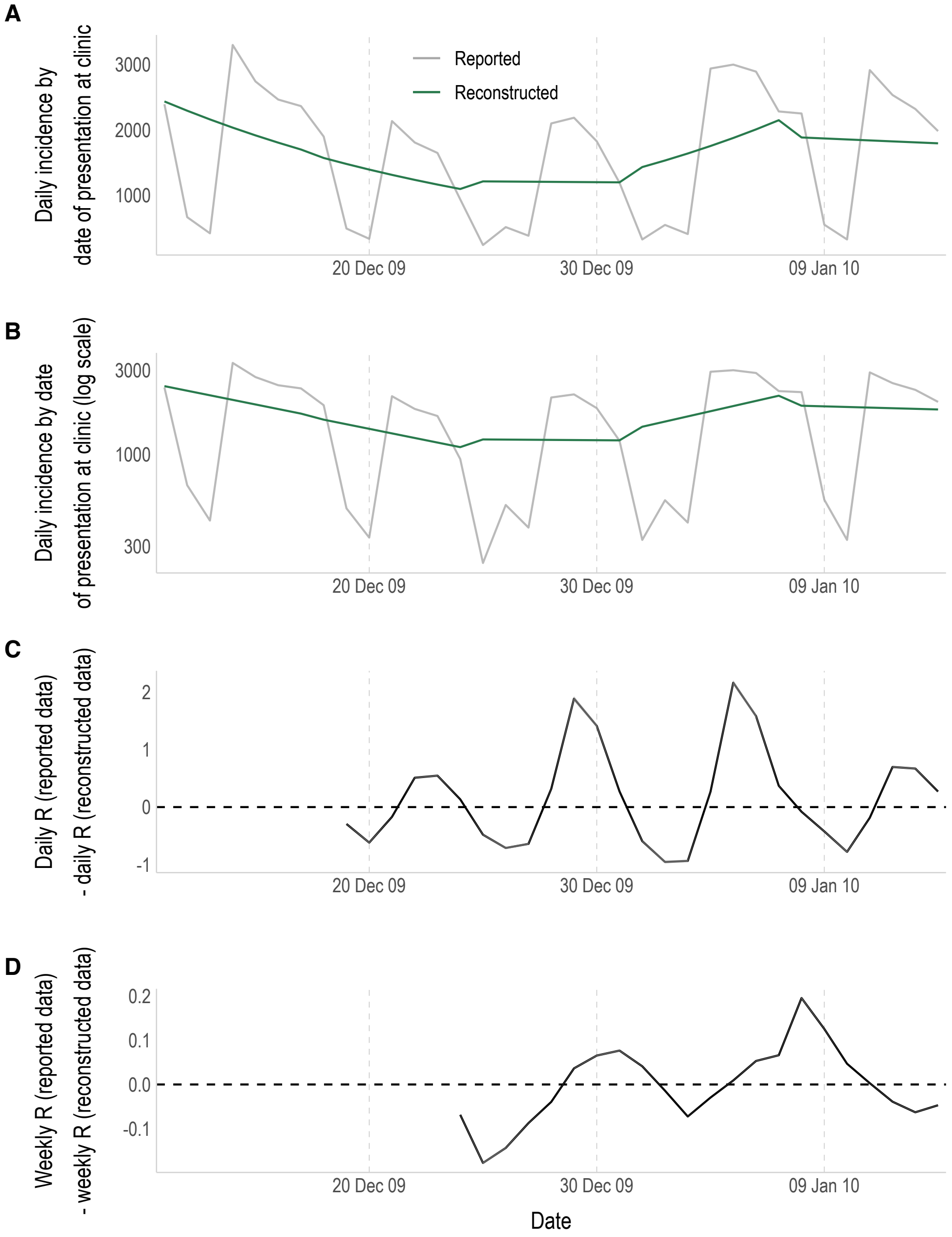

**Figure S2.** A-B) The reported (grey) and reconstructed (green) daily incidence of influenza by the date of presentation at the military clinic, either on a natural scale (A) or log scale (B). C-D) The absolute difference in the C) daily and D) weekly sliding $R_t$ estimates made using the reported daily data and the reconstructed daily data. Note that the y-axis scale is different in panels C and D.

#### b. COVID-19 cases

In the first case study for COVID-19, the overall trends in $R_t$ over time using the reported and reconstructed daily data were similar for both the daily and weekly sliding $R_t$ estimates (Figure S3). The daily $R_t$ estimates using the reconstructed data considerably smoothed out the impact of weekend effects, which were prominent in the reported incidence (Figure S3A, Figure S4A-B). The weekly sliding $R_t$ estimates are very similar using both datasets, with some minor discrepancies e.g., around early September and mid-December 2020 (Figure S3B).

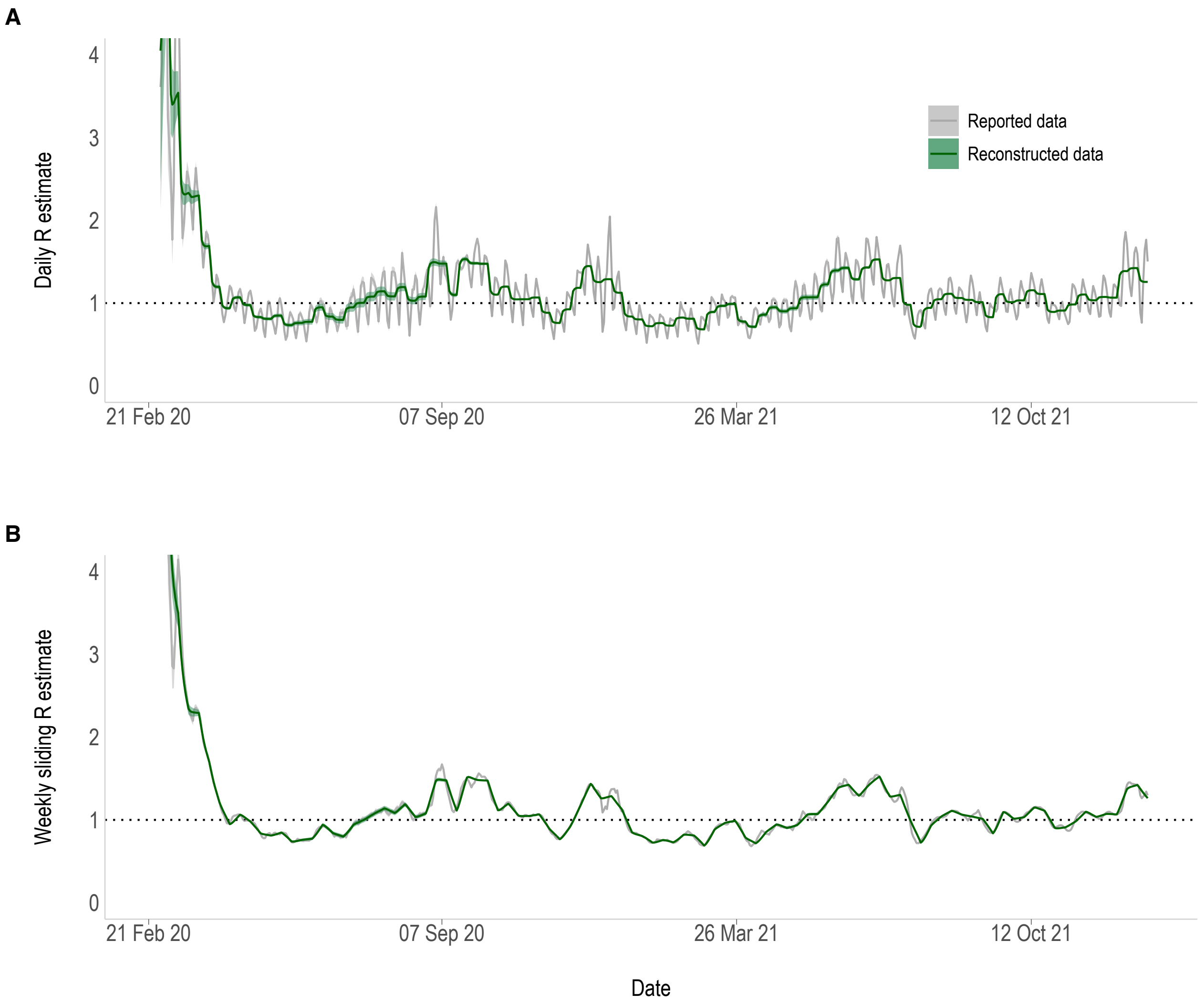

**Figure S3.** The A) daily and B) weekly sliding $R_t$ estimates for COVID-19 cases based on the reported (grey) and the reconstructed (green) daily data by date of specimen. $R_t$ estimates start on the first day of the second aggregation window (day 8 – 28th February 2020) and are plotted at the end of the time window. Shading corresponds to the 95% credible interval of the estimates. The y-axis has been cropped to a maximum of 4 for clarity.

When comparing the absolute difference in the $R_t$ estimates made using the reported and reconstructed data, the larger spikes coincide with periods of either low incidence or more prominent weekend effects in the reported data (Figure S4).

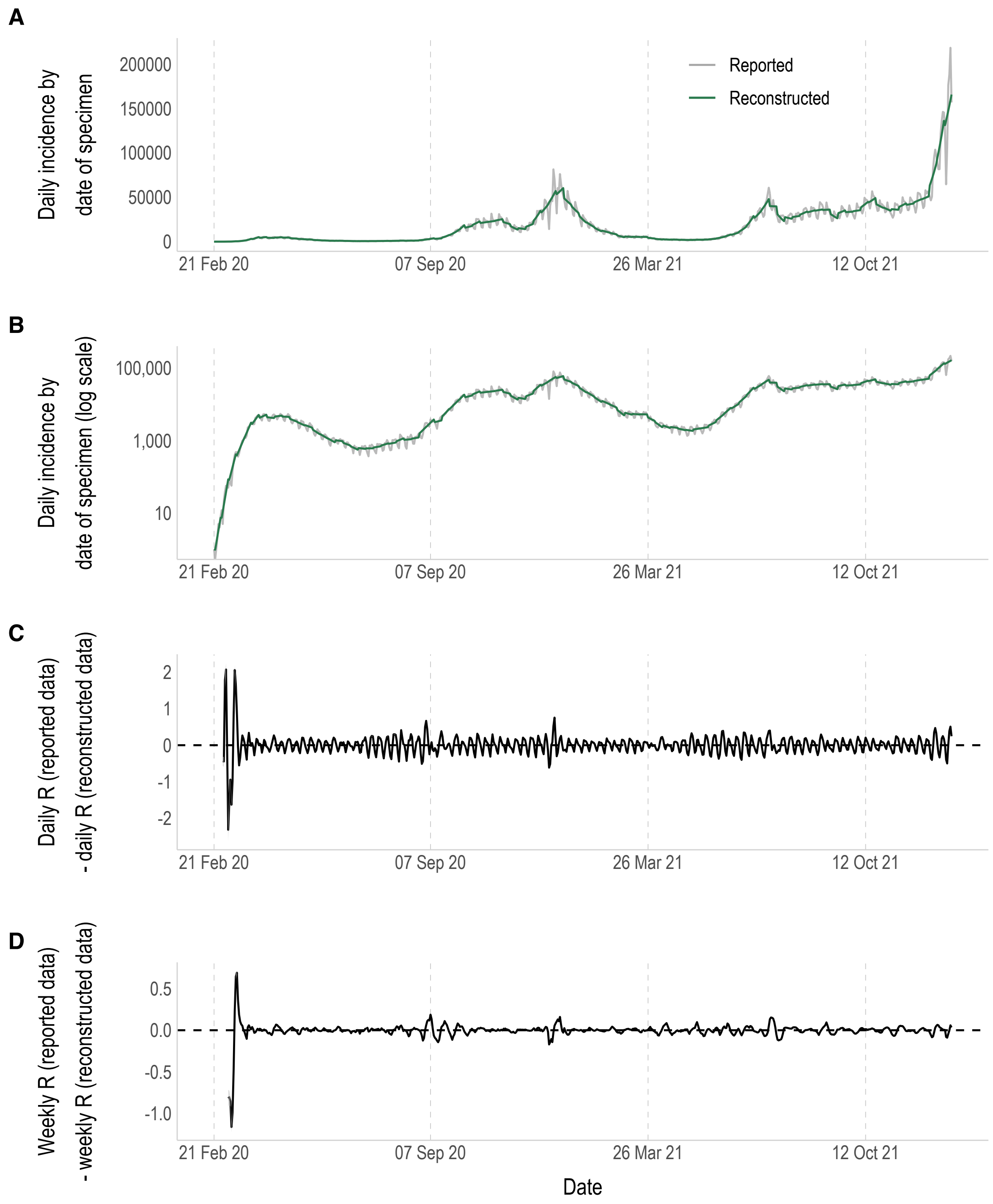

**Figure S4.** A-B) The reported (grey) and reconstructed (green) daily incidence of COVID-19 by the date of specimen, either on a natural scale (A) or log scale (B). C-D) The absolute difference in the C) daily and D) weekly sliding $R_t$ estimates made using the reported data and the reconstructed data. Note that the y-axis scale is different in panels C and D.

#### c. COVID-19 deaths

In the second case study for COVID-19, due to the lower incidence of COVID-19 deaths compared to COVID-19 cases, there is greater uncertainty in the $R_t$ estimates (Figure S5). There is still some indication of noise in this dataset, but the pattern appears less pronounced and more irregular than the variation caused by weekend effects as seen in the COVID-19 cases data. Unlike the data for flu and COVID-19 cases, further analysis showed that there was no pattern in reporting depending on the day of the week (Figure S7).

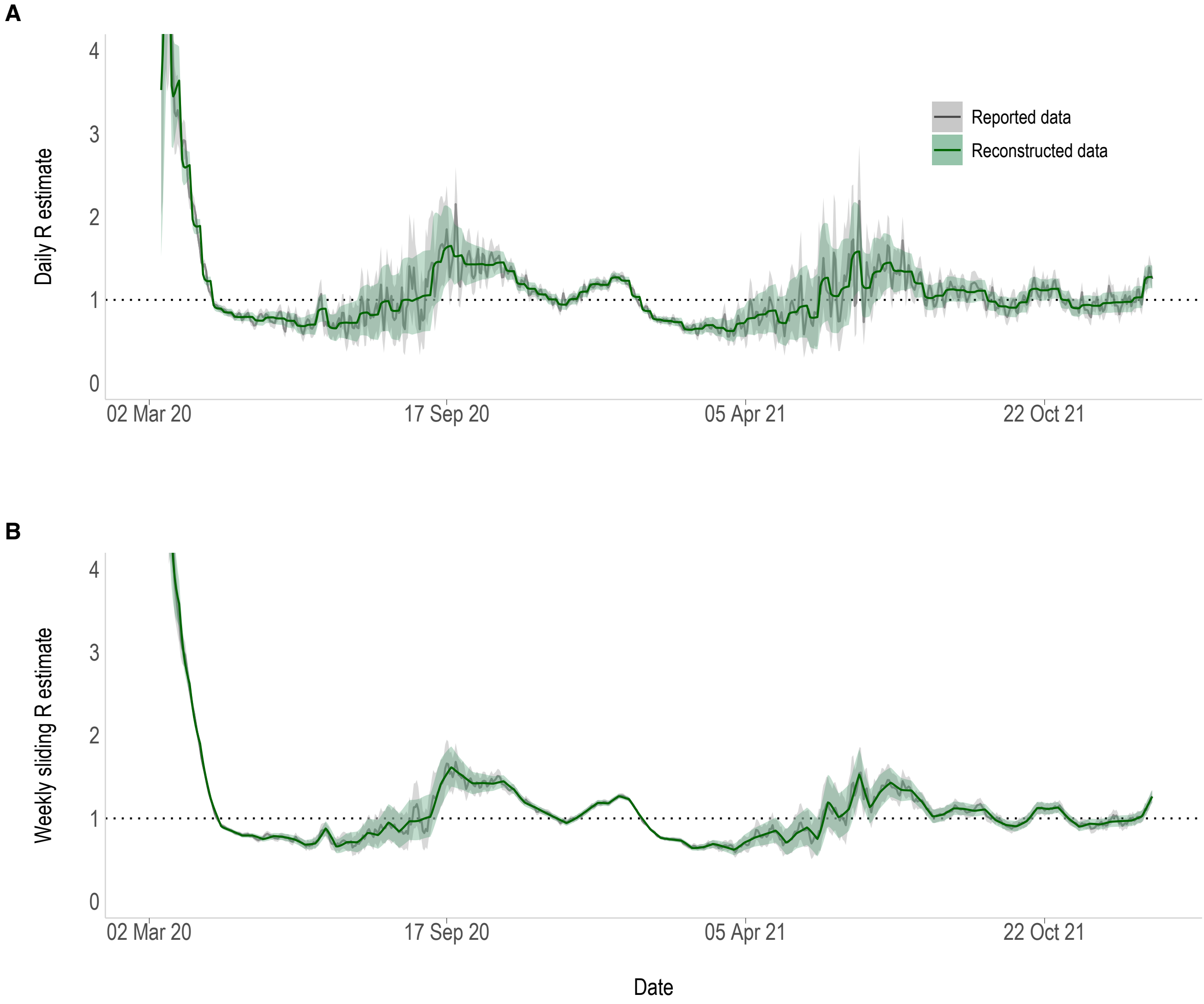

**Figure S5.** The A) daily and B) weekly sliding $R_t$ estimates for COVID-19 deaths based on the reported daily data (grey) and the reconstructed daily data (green) by date of death within 28 days of a positive test. $R_t$ estimates start on the first day of the second aggregation window (day 8 – 9$^{th}$ March 2020) and are plotted at the end of the time window. Shading corresponds to the 95% credible interval of the estimates. The y-axis has been cropped to a maximum of 4 for clarity.

In contrast with COVID-19 case data, the greatest differences in $R_t$ estimates obtained from death data appear to coincide solely with periods of low incidence (Figure S6).

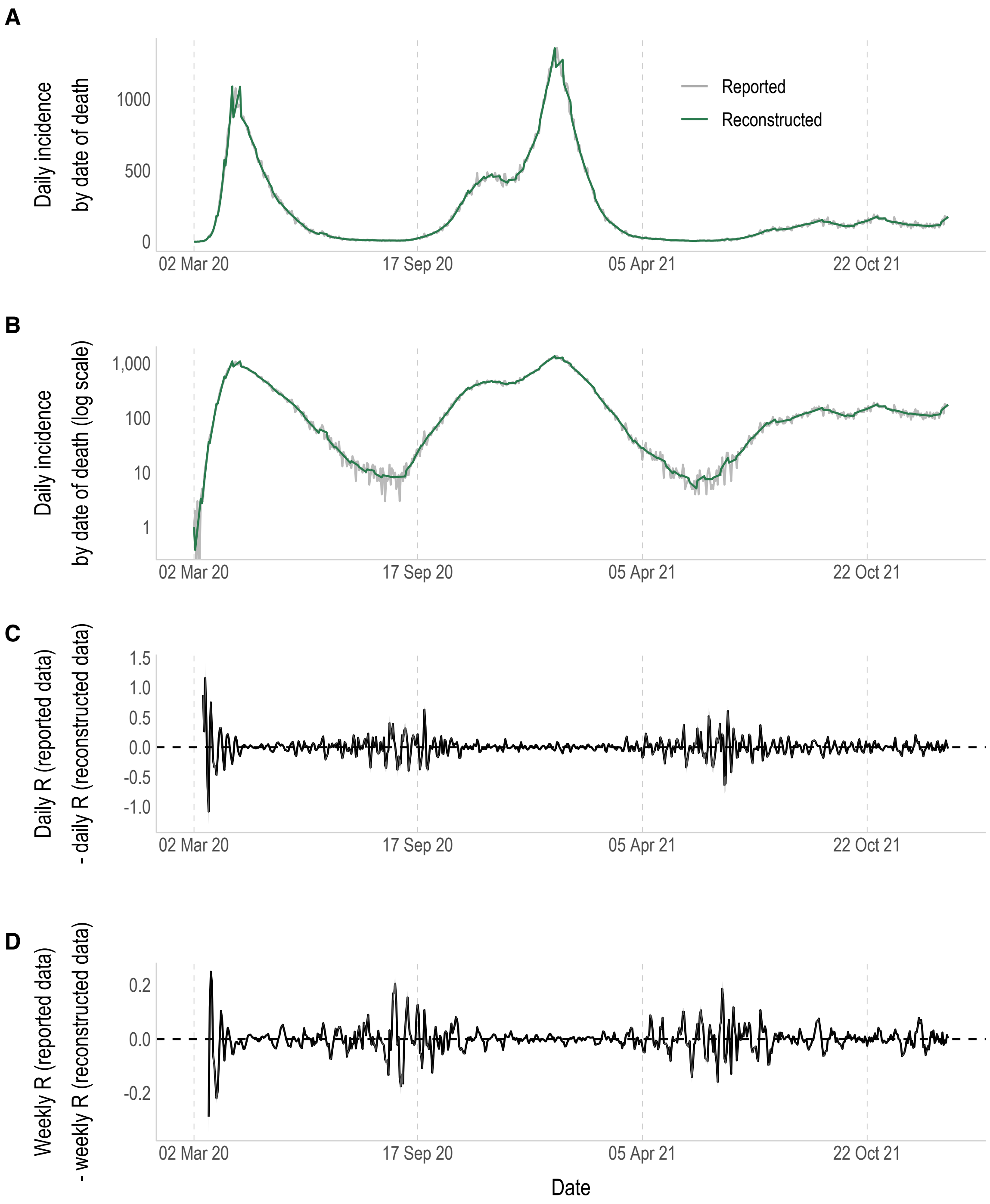

**Figure S6.** A-B) The reported (grey) and reconstructed (green) daily incidence of COVID-19 deaths within 28 days of a positive test, either on a natural scale (A) or log scale (B). C-D) The absolute difference in the C) daily and D) weekly sliding $R_t$ estimates made using the reported data and the reconstructed data. Note that the y-axis scale is different in panels C and D.

#### d. Reported incidence patterns by weekday

There is a clear pattern in the reporting of influenza and COVID-19 cases, with generally higher incidence reported on Mondays, which then declines throughout the week, with the lowest incidence on the weekends (Figure S7A-B). Reported incidence appears lower on Fridays for influenza, however this dataset encompasses Christmas Day 2009 and New Year's Day 2010, which both fell on Friday. On the other hand, COVID-19 deaths did not appear to show any pattern in reporting, with no clear deviations from the weekly mean incidence regardless of the weekday (Figure S7C).

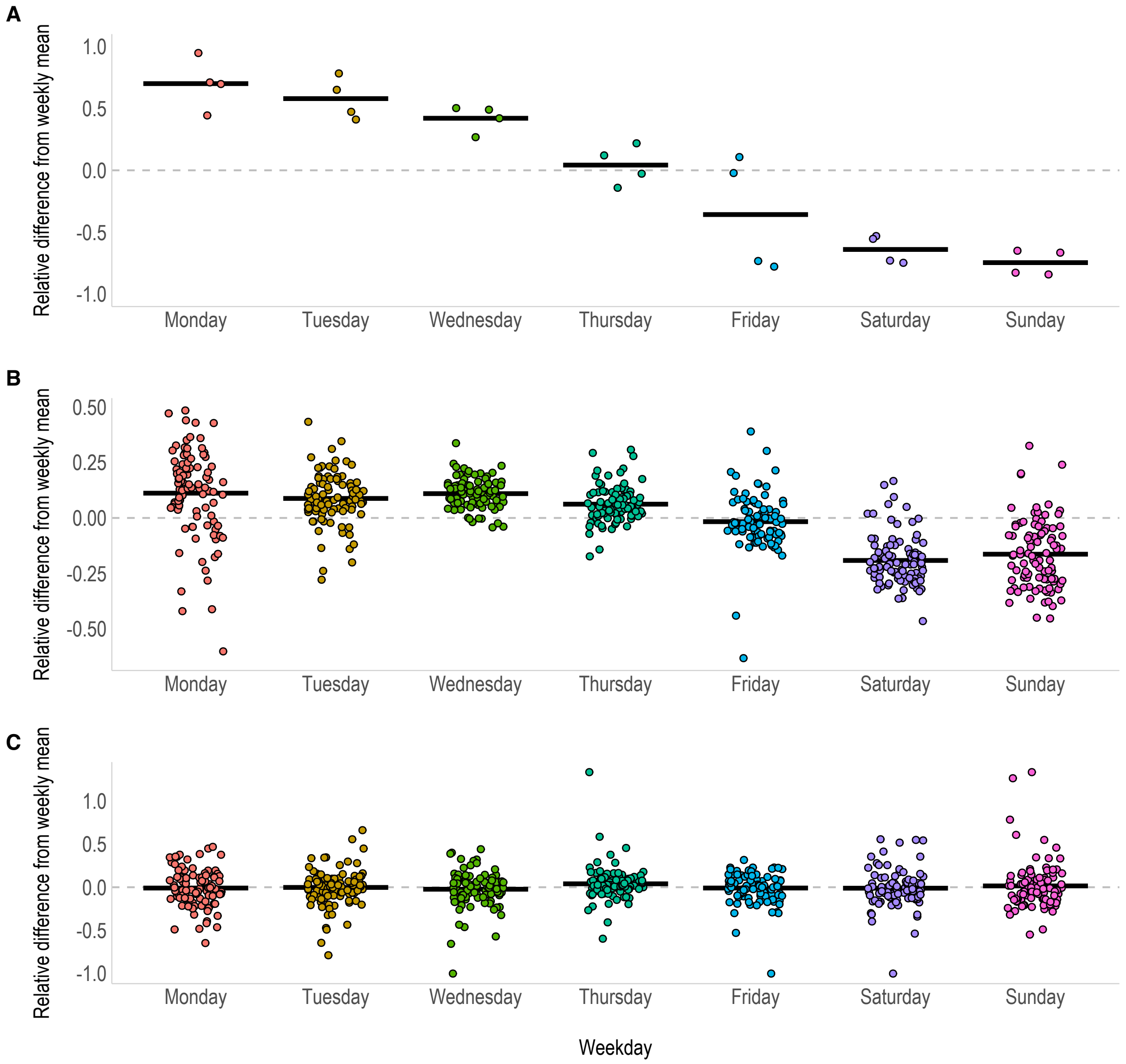

Figure S7. The relative difference between the reported incidence for A) influenza, B) COVID-19 cases and C) COVID-19 deaths, on each weekday compared to the mean incidence for that week. Only data for full calendar weeks are included. Black lines represent the mean relative difference and the dashed grey line indicates zero (corresponding to no difference between the reported incidence on that day compared to the weekly mean).

#### e. Classification tables

We directly compared the classification of the epidemic as increasing (95% credible interval of the $R_t$ estimate is above 1), uncertain (95% credible interval encompasses 1), and declining (95% credible interval is below 1), depending on whether the $R_t$ estimates were made using the reported or reconstructed daily data (Figure S8). The overall agreement in the classification of daily $R_t$ estimates for influenza, COVID-19 cases, and COVID-19 deaths, was 44.4%, 74.4%, and 85.8% respectively. The overall agreement in the classification of weekly sliding $R_t$ estimates was higher for each, with 81.8%, 94.9% and 93.3% agreement respectively. There is a greater correlation between the classification of estimates made using COVID-19 death data, reflected by the dark blue diagonal line across the grid squares. This is likely due to the reported deaths being less affected by weekend effects (see section 2d), and therefore the reported and reconstructed data (and the $R_t$ estimates made from them) are more similar (Figure S8E-F).

0% | 1-15% | 16-30% | 31-45% | 46-60% | 61-75% | 76-90% | 91-100%

*Daily R* | *Weekly sliding R*

*Influenza*

A

| | | 95% CrI for estimates using **reported** daily data (*n=27*) | | |
|---|---|---|---|---|
| | | Above 1 | Encompassing 1 | Below 1 |
| Agreement of the 95% CrI for estimates using **reconstructed** daily data (*n=27*) | Above 1 | 43.8% (*n=7*) | 0% (*n=0*) | 27.3% (*n=3*) |
| | Encompassing 1 | 25% (*n=4*) | 0% (*n=0*) | 27.3% (*n=3*) |
| | Below 1 | 31.3% (*n=5*) | 0% (*n=0*) | 45.5% (*n=5*) |
| *Total (n)* | | *16* | *0* | *11* |

B

| | | 95% CrI for estimates using **reported** daily data (*n=22*) | | |
|---|---|---|---|---|
| | | Above 1 | Encompassing 1 | Below 1 |
| Agreement of the 95% CrI for estimates using **reconstructed** daily data (*n=22*) | Above 1 | 77.8% (*n=7*) | 100% (*n=1*) | 0% (*n=0*) |
| | Encompassing 1 | 11.1% (*n=1*) | 0% (*n=0*) | 8.3% (*n=1*) |
| | Below 1 | 11.1% (*n=1*) | 0% (*n=0*) | 91.7% (*n=11*) |
| *Total (n)* | | *9* | *1* | *12* |

*COVID-19 cases*

C

| | | 95% CrI for estimates using **reported** daily data (*n=650*) | | |
|---|---|---|---|---|
| | | Above 1 | Encompassing 1 | Below 1 |
| Agreement of the 95% CrI for estimates using **reconstructed** daily data (*n=650*) | Above 1 | 80.7% (*n=205*) | 35.5% (*n=11*) | 21.9% (*n=80*) |
| | Encompassing 1 | 5.5% (*n=14*) | 12.9% (*n=4*) | 2.7% (*n=10*) |
| | Below 1 | 13.8% (*n=35*) | 51.6% (*n=16*) | 75.3% (*n=275*) |
| *Total (n)* | | *254* | *31* | *365* |

D

| | | 95% CrI for estimates using **reported** daily data (*n=650*) | | |
|---|---|---|---|---|
| | | Above 1 | Encompassing 1 | Below 1 |
| Agreement of the 95% CrI for estimates using **reconstructed** daily data (*n=650*) | Above 1 | 96.2% (*n=255*) | 40% (*n=8*) | 2.5% (*n=9*) |
| | Encompassing 1 | 1.5% (*n=4*) | 35% (*n=7*) | 0.3% (*n=1*) |
| | Below 1 | 2.3% (*n=6*) | 25% (*n=5*) | 97.3% (*n=355*) |
| *Total (n)* | | *265* | *20* | *365* |

*COVID-19 deaths*

E

| | | 95% CrI for estimates using **reported** daily data (*n=643*) | | |
|---|---|---|---|---|
| | | Above 1 | Encompassing 1 | Below 1 |
| Agreement of the 95% CrI for estimates using **reconstructed** daily data (*n=643*) | Above 1 | 81.8% (*n=157*) | 6.7% (*n=18*) | 0% (*n=0*) |
| | Encompassing 1 | 18.2% (*n=35*) | 91.1% (*n=246*) | 17.7% (*n=32*) |
| | Below 1 | 0% (*n=0*) | 2.2% (*n=6*) | 82.3% (*n=149*) |
| *Total (n)* | | *192* | *270* | *181* |

F

| | | 95% CrI for estimates using **reported** daily data (*n=643*) | | |
|---|---|---|---|---|
| | | Above 1 | Encompassing 1 | Below 1 |
| Agreement of the 95% CrI for estimates using **reconstructed** daily data (*n=643*) | Above 1 | 98.3% (*n=226*) | 3.5% (*n=6*) | 0% (*n=0*) |
| | Encompassing 1 | 1.7% (*n=4*) | 86.0% (*n=147*) | 6.2% (*n=15*) |
| | Below 1 | 0% (*n=0*) | 10.5% (*n=18*) | 93.8% (*n=227*) |
| *Total (n)* | | *230* | *171* | *242* |

**Figure S8.** Comparison of the percentage agreement in the classification of $R_t$ estimates as above 1 (lower bound of the 95% CrI is above 1), encompassing 1, or below 1 (upper bound of the 95% CrI is below 1), when using reported and reconstructed incidence data. Daily and weekly sliding $R_t$ estimates are compared for influenza cases (A-B), COVID-19 cases (C-D), and COVID-19 deaths (E-F). Due to low incidence, the first 30 days of data is excluded for both COVID-19 datasets. Darker shades of blue correspond to greater percentage agreement, with a strong correlation represented by a dark diagonal line across the grid squares from top left to bottom right.

#### f. Computational time

All real data scenarios took less than 3 seconds to run on MacOS (2 GHz Quad-Core Intel Core i5) 16GB RAM (Table S1). The influenza cases scenario, with >57,000 cases, took 2 seconds to run. The COVID-19 cases and deaths scenarios, which estimated $R_t$ over 97 and 96 weeks of incidence data, took 3 seconds to run.

**Table S1.** Time taken to estimate $R_t$ in each of the real data scenarios.

| Real data scenario | Length of time period analysed | Total incidence | Estimation time |
|---|---|---|---|
| Influenza cases | 5 weeks | 57,351 | 2s |
| COVID-19 cases | 97 weeks | 13,139,522 | 3s |
| COVID-19 deaths | 96 weeks | 149,557 | 3s |

### 3. Simulation study

The simulation study involved assessing the performance of the method in multiple epidemic contexts. These included scenarios where $R_t$ remains constant over time, or where $R_t$ varies over time, with either a sudden stepwise change or a gradual change. For each scenario, 100 epidemic trajectories were stochastically generated using the R package projections.[3] Each epidemic was seeded with 10 days of 7 daily cases and then simulated over 70 days (10 weeks). The values of $R_t$ estimated from the simulated incidence data were evaluated in terms of their bias, uncertainty and 95% coverage (Table S2).

**Table S2.** Definitions for the criteria used to assess the performance of our method in the simulation study.

| Criterion | Definition |
|---|---|
| Bias | Absolute difference between the mean $R_t$ estimate across the 100 simulations and the true value of $R_t$ that the simulated incidence is based on. |
| Uncertainty | The mean width (across the 100 simulations) of the 95% credible interval for $R_t$ estimates. |
| 95% coverage | The proportion of $R_t$ estimates across the 100 simulations with the true value of $R_t$ within the 95% credible interval. |

#### a. Constant $R_t$

For a scenario where $R_t$ remains constant over time, four values of $R_t$ were considered: 1, 1.25, 1.5 and 1.75. The $R_t$ estimates all recovered the true value that the simulations were based on, with very little difference between the estimates made using daily and weekly data (Figure S9A-D). There was no evidence of bias in the estimates (Figure S9E-H), and as expected, the uncertainty declined over time as the number of cases rose, and remained the same when $R_t$ was 1, reflecting stable case numbers (Figure S9I-L). The 95% coverage was consistently high (Figure S9M-P) and on average the true value of $R_t$ was encompassed by the 95% credible interval 96-97% of the time in each scenario. However, the wavy pattern in these plots is likely to correspond to discontinuities in the reconstructed incidence data (section 3b, Figure S10).

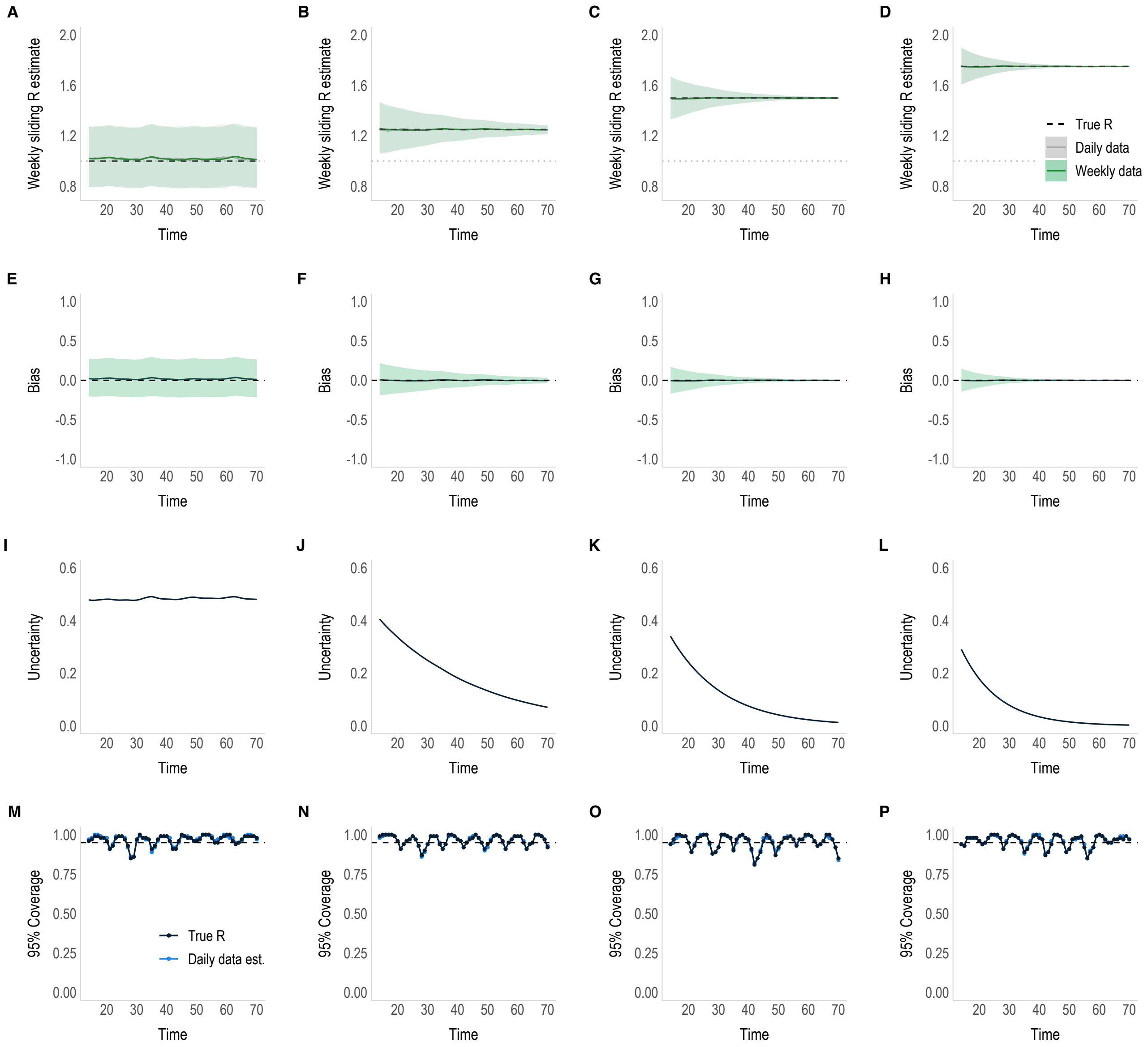

**Figure S9.** Performance of the method when estimating a constant $R_t$ for 100 simulated epidemics. A-D) The mean weekly sliding $R_t$ estimates using daily data (grey) and weekly data (green). The black dashed line represents the true value of $R_t$ that the 100 simulations are based on, which is either A) 1, B) 1.25, C) 1.5, or D) 1.75. The estimates and their 95% credible intervals (shaded areas) are very similar and therefore overlap. E-H) The bias (or absolute difference) in the mean $R_t$ estimated from weekly data compared to the true value of $R_t$. The shaded area corresponds to the 95% quantiles of the bias across the 100 simulations. I-L) The uncertainty (mean width of the 95% credible interval) in the $R_t$ values estimated from weekly data. M-P) The proportion of the 100 $R_t$ estimates obtained from weekly data where the 95% credible interval encompasses either the true value of $R_t$ (black) or the value of $R_t$ that would have been estimated from daily data (blue). Here, the 95% coverage is similar for both the true value of $R_t$ and the daily data estimate, and therefore they overlap.

#### b. Discontinuities in the reconstructed incidence data

As part of the process of reconstructing daily incidence data from aggregated data, the reconstructed incidence is slightly adjusted using a constant ($k_w$) to ensure that if you were to re-aggregate it, it would still match the original aggregated data used as the input. This can result in discontinuities in the borders between time periods that the data has been aggregated over, which means that the reconstructed incidence is not completely smooth, even when there is no noise in the data (Figure S10). When $R_t$ is estimated using sliding windows, these may align perfectly with the time windows that data were aggregated over or they may encompass the border between aggregations, and therefore the discontinuity in the data. As shown in Figure S9M-P, this can lead to a wavy pattern in the 95% coverage, where the sliding window moves across the incidence data and can be more/less affected by the discontinuities depending on where they fall. This is important to be aware of, but as seen in the real data scenarios (section 1), reported daily data is often dramatically affected by intra-weekly variations in reporting, such as weekend effects. The reconstructed data, even with these discontinuities, is much smoother than the daily data that is typically reported. Nevertheless, we suggest that the sliding time window used to estimate $R_t$ should be equal to or longer than the length of the aggregation window, to limit the effect of the discontinuities on the estimates.

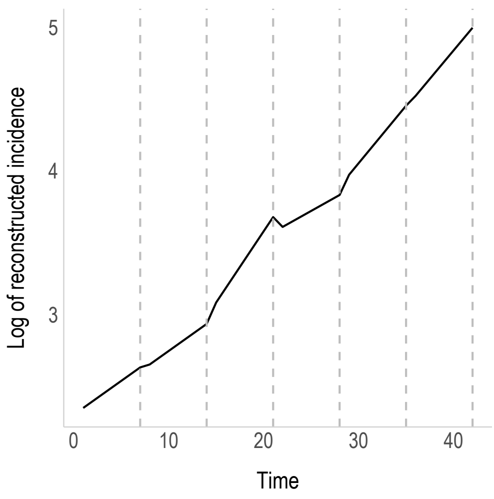

**Figure S10.** An example of reconstructed daily incidence data (on the log scale) with clear discontinuities on the borders between time periods where the data had been aggregated (grey dashed lines).

#### c. Time varying $R_t$: sudden change

In the first scenario for time varying transmissibility, we considered a stepwise decrease or increase in the value of $R_t$. $R_t$ may suddenly decrease or increase following the rapid implementation or relaxation of stringent control measures, for instance, a strict mass 'lockdown' event.

We considered four decreasing scenarios, where on day 35 of the simulated outbreak, $R_t$ falls from: 1.25 to 0.75, 1.25 to 1, 1.5 to 1.25, or 1.75 to 1.5. The true value of $R_t$ was successfully recovered in all scenarios, except for a week-long delay following the step change (Figure S11A-D), which corresponds to dips in the bias plots (Figure S11E-H). This delay is due to the weekly sliding window used for $R_t$ estimation, which would encompass incidence data before and after the step change.

As expected, uncertainty declines as case numbers rise when $R_t > 1$ (Figure S11I-L). The decrease in $R_t$ from 1.25 to 0.75 replicates a situation where control measures successfully bring $R_t$ below 1, resulting in falling case numbers and increased uncertainty after day 35 (Figure S11I). Similarly, when $R_t$ falls from 1.25 to 1, case numbers become stable, leading to a plateau in the uncertainty (Figure S11J).

Importantly, despite not recovering the true value of $R_t$ in the week following the step change, a high proportion of the estimates recovered the value of $R_t$ that would have been estimated from the reported daily data (Figure S11M-P). There is a slightly larger drop in the 95% coverage when $R_t$ falls from 1.75 to 1.5 (Figure S11P), however, this is due to higher case numbers in this scenario resulting in very narrow 95% credible intervals. Overall, the 95% credible interval encompassed the true value of $R_t$ 85-90% of the time, which rose to 95-97% if the week following the step change was excluded.

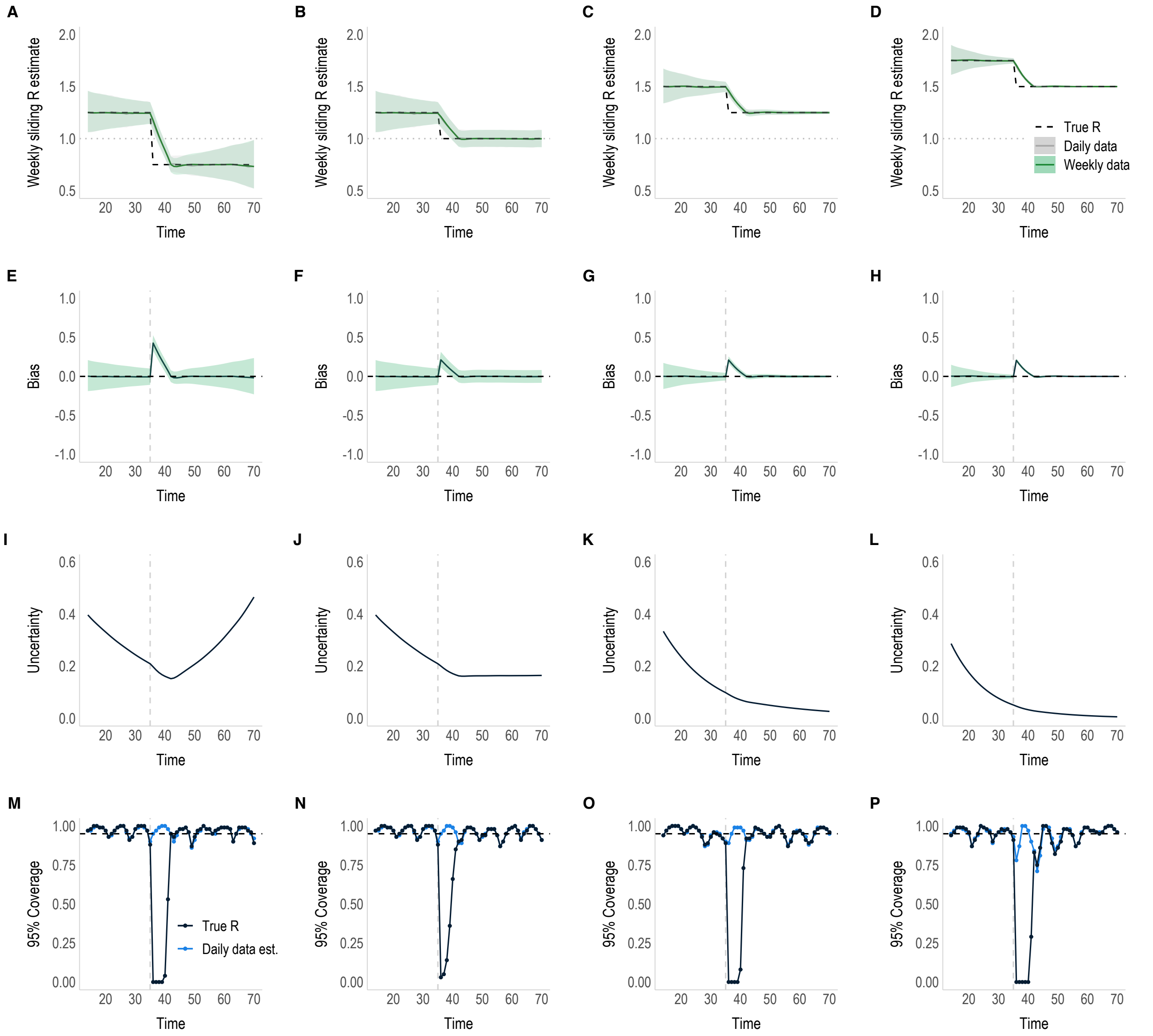

**Figure S11.** Performance of the method when estimating a time varying $R_t$ for 100 simulated epidemics. In this scenario, $R_t$ suddenly decreases on day 35 (grey dashed line) and remains constant before and after the step change. A-D) The mean weekly sliding $R_t$ estimates using daily data (grey) and weekly data (green). The black dashed line represents the true value of $R_t$ that the 100 simulations are based on, which is either a decrease from A) 1.25 to 0.75, B) 1.25 to 1, C) 1.5 to 1.25, or D) 1.75 to 1.5. The grey dotted line represents the threshold of $R_t = 1$. The estimates (plotted at the end of each time window) and their 95% credible intervals (shaded area) are very similar and therefore overlap. E-H) The bias (or absolute difference) in the mean $R_t$ estimated from weekly data compared to the true value of $R_t$. The shaded area corresponds to the 95% quantiles of the bias across the 100 simulations. I-L) The uncertainty (mean width of the 95% credible interval) in the $R_t$ values estimated from weekly data. M-P) The proportion of the 100 $R_t$ estimates made using weekly data where the 95% credible interval encompasses either the true value of $R_t$ (black) or the value of $R_t$ that would have been estimated from daily data (blue).

A sudden stepwise increase in $R_t$ is perhaps more difficult to explain in a real-world context, but it is also shown here for the sake of completion. Three scenarios for a stepwise increase in $R_t$ were considered: 1 to 1.25, 1.25 to 1.5, and 1.5 to 1.75. Similarly, the true value of $R_t$ was well recovered in each scenario, but there was a week-long delay following the step change (Figure S12A-F). The uncertainty was stable when $R_t$ is 1 and falls as case numbers rise (Figure S12G-I). Finally, despite the drop in 95% coverage for the true value of $R_t$, the weekly estimates recovered what would have been estimated from daily data (Figure S12J-L). Across these scenarios, the 95% credible interval encompassed the true value of $R_t$ 87-95% of the time, rising to 95-96% if the week following the step change is excluded.

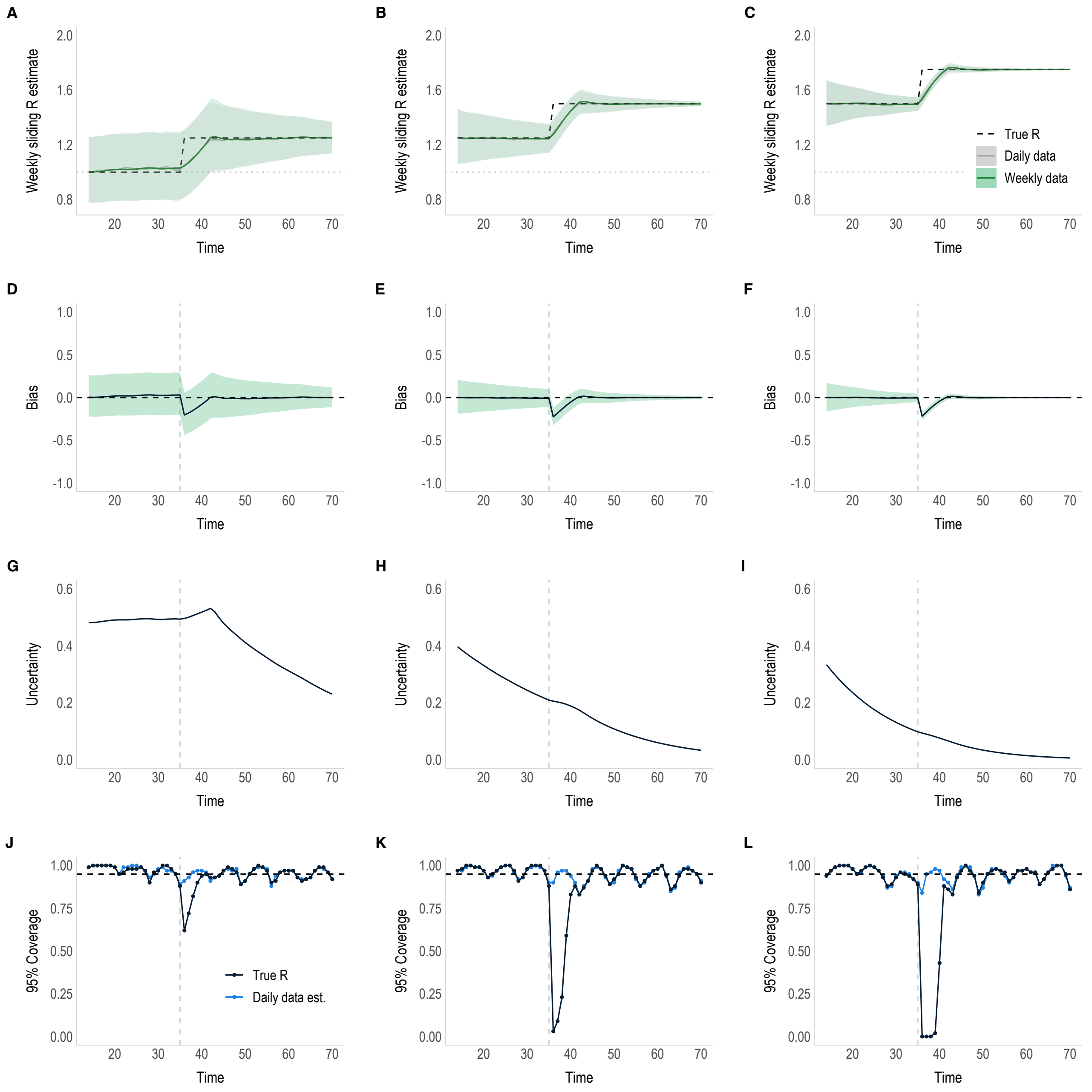

**Figure S12.** Performance of the method when estimating a time varying $R_t$ for 100 simulated epidemics. In this scenario, $R_t$ suddenly increases on day 35 (grey dashed line) and remains constant before and after the step change. A-C) The mean weekly sliding $R_t$ estimates using daily data (grey) and weekly data (green). The black dashed line represents the true value of $R_t$ that the 100 simulations are based on, which is either an increase from A) 1 to 1.25, B) 1.25 to 1.5, or C) 1.5 to 1.75. The grey dotted line represents the threshold of $R_t = 1$. The estimates (plotted at the end of each time window) and their 95% credible intervals (shaded area) are very similar and therefore overlap. D-F) The bias (or absolute difference) in the mean $R_t$ estimated from weekly data compared to the true value of $R_t$. The shaded area corresponds to the 95% quantiles of the bias across the 100 simulations. G-I) The uncertainty (mean width of the 95% credible interval) in the $R_t$ values estimated from weekly data. J-L) The proportion of the 100 $R_t$ estimates made using weekly data where the 95% credible interval encompasses either the true value of $R_t$ (black) or the value of $R_t$ that would have been estimated from daily data (blue).

#### d. Time-varying $R_t$: gradual change

Changes in transmissibility are often more gradual over time. For example, $R_t$ could slowly decrease as a population becomes more aware of a circulating pathogen and modifies their behaviour accordingly, or $R_t$ could increase in response to the gradual easing of restrictions, such as social distancing measures, or a gradual decline in compliance to such restrictions. In the following scenarios, we considered a gradual change in $R_t$ that occurred over the course of 30 days, starting on day 20 and ending on day 50. Four scenarios were considered for a gradually decreasing $R_t$, where $R_t$ falls from: 1.25 to 0.75, 1.25 to 1, 1.5 to 1.25, and 1.75 to 1.5 (Figure S13). Three scenarios were considered for a gradually increasing $R_t$, where $R_t$ increases from: 1 to 1.25, 1.25 to 1.5, and 1.5 to 1.75.

As above, during the period of gradual change, the estimates were affected by the lag due to the weekly sliding window for $R_t$ estimation (Figure S13A-D & Figure S14A-C). This means that $R_t$ was slightly overestimated during the 30 days that $R_t$ was gradually decreasing and slightly underestimated when $R_t$ was gradually increasing (Figure S13E-H & Figure S14D-F). The uncertainty declined as case numbers rose (Figures S13I-L & S14G-I), increased when $R_t$ fell below 1 (Figure S13I) and plateaued when $R_t = 1$ (Figures S13J & S14G). The lag in the estimates caused some notable drops in the 95% coverage (Figures S13M-P & S14J-L). This is particularly prominent when incidence is high and credible intervals are small, such as in the gradual decrease in $R_t$ from 1.75 to 1.5 scenario, where the true value of $R_t$ was encompassed by the 95% credible interval 62% of the time (Figure S13P). For all other gradually decreasing scenarios and the gradually increasing scenarios, true $R_t$ was recovered between 81-94% and 78-96% of the time respectively. It is important to note, however, that even when substantial dips in coverage occurred, the bias remained small, meaning there was very little difference between the estimate and the true value of $R_t$ despite not being exactly the same (Figure S13E-H).

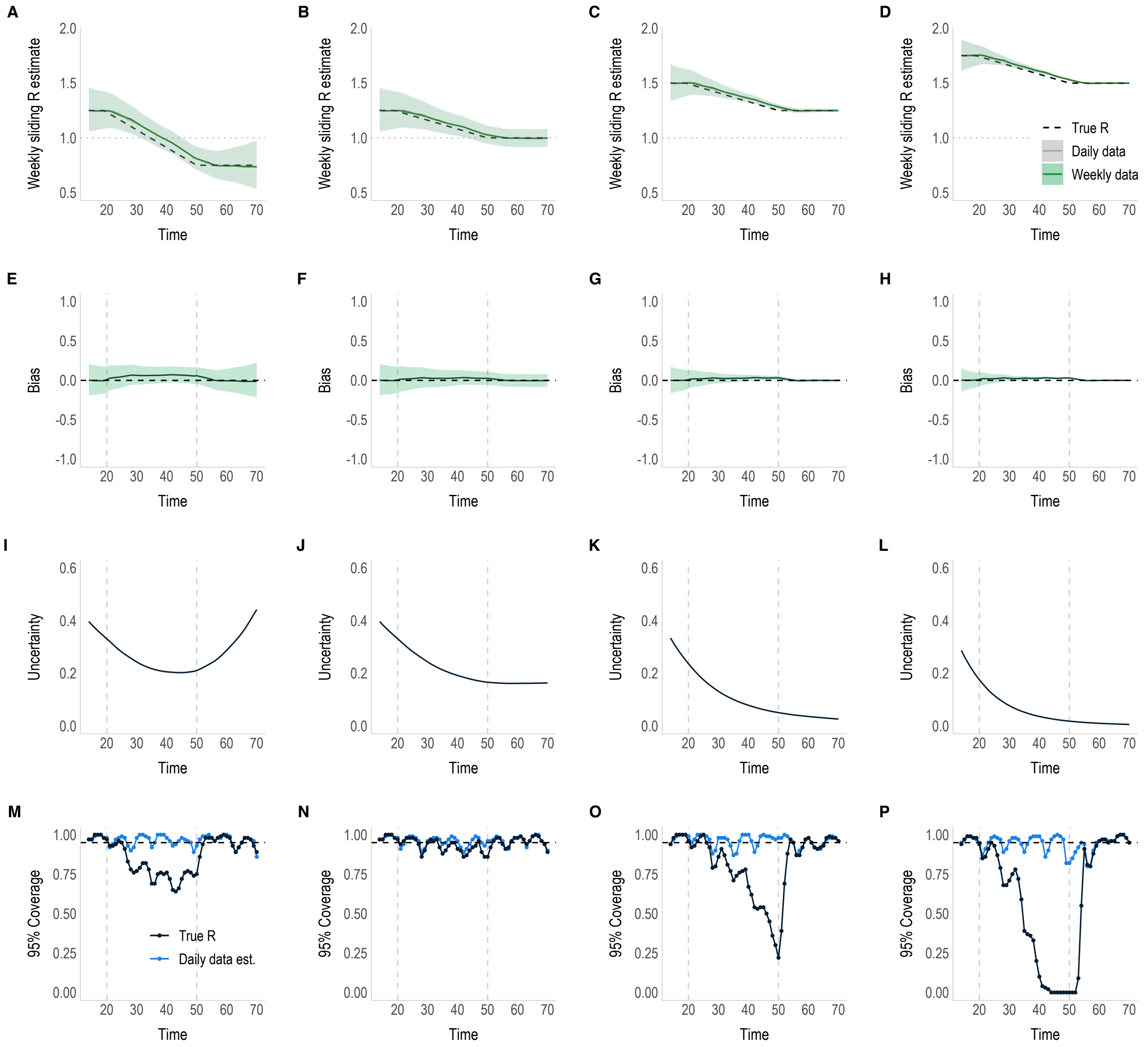

**Figure S13.** Performance of the method when estimating a time varying $R_t$ for 100 simulated epidemics. In this scenario, $R_t$ gradually decreases over the course of 30 days (day 20 to day 50, shown with grey dashed lines), and remains constant before and after the change. A-D) The mean weekly sliding $R_t$ estimates using daily data (grey) and weekly data (green). The black dashed line represents the true value of $R_t$ that the 100 simulations are based on, which is either a decline from A) 1.25 to 0.75, B) 1.25 to 1, C) 1.5 to 1.25, or D) 1.75 to 1.5. The grey dotted line represents the threshold of $R_t = 1$. The estimates (plotted at the end of each time window) and their 95% credible intervals (shaded area) are very similar and therefore overlap. E-H) The bias (or absolute difference) in the mean $R_t$ estimated from weekly data compared to the true value of $R_t$. The shaded area corresponds to the 95% quantiles of the bias across the 100 simulations. I-L) The uncertainty (mean width of the 95% credible interval) in the $R_t$ values estimated from weekly data. M-P) The proportion of the 100 $R_t$ estimates made using weekly data where the 95% credible interval encompasses either the true value of $R_t$ (black) or the value of $R_t$ that would have been estimated from daily data (blue).

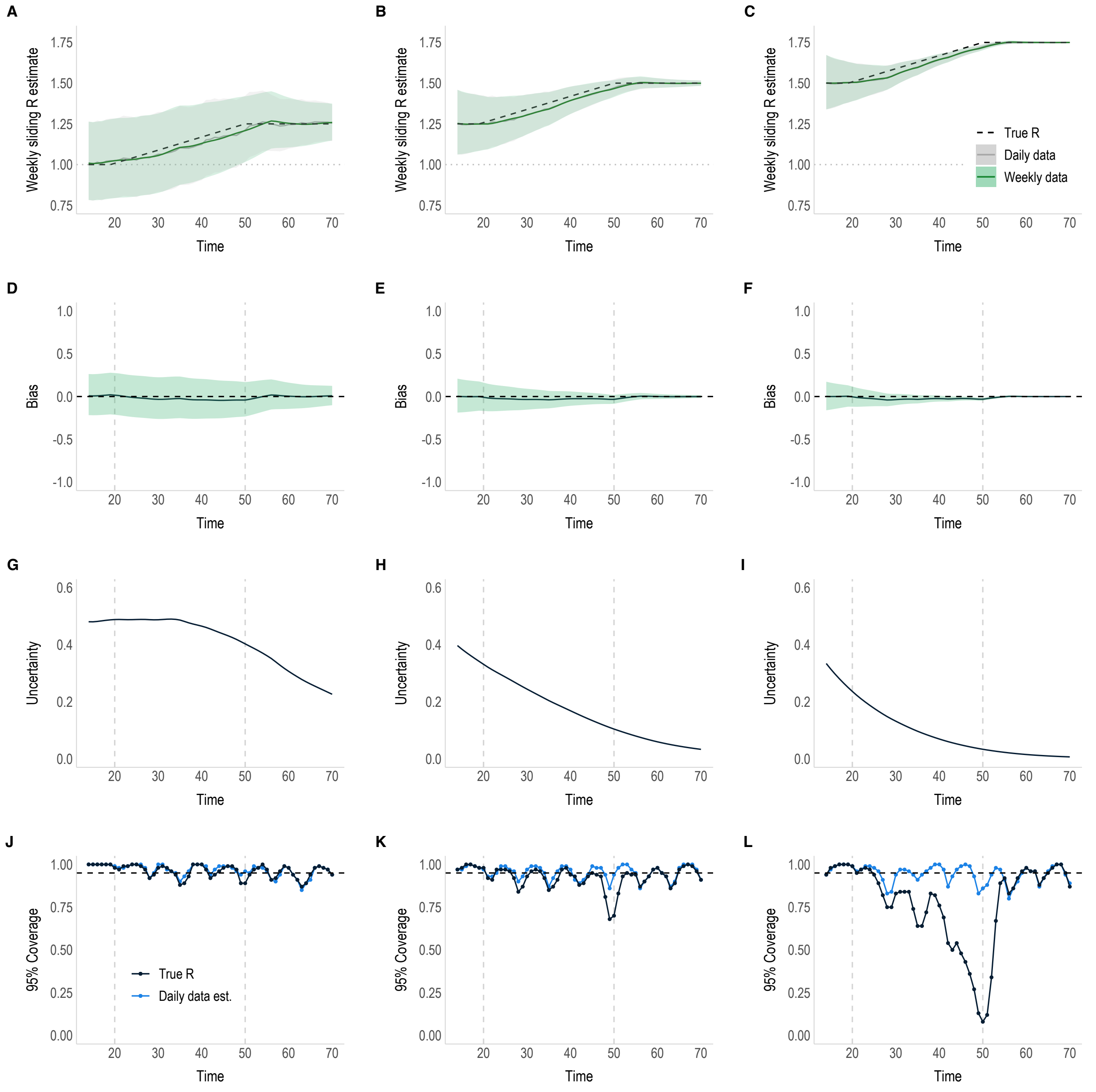

**Figure S14.** Performance of the method when estimating a time varying $R_t$ for 100 simulated epidemics. In this scenario, $R_t$ gradually increases over the course of 30 days (day 20 to day 50, shown with grey dashed lines), and remains constant before and after the change. A-C) The mean weekly sliding $R_t$ estimates using daily data (grey) and weekly data (green). The black dashed line represents the true value of $R_t$ that the 100 simulations are based on, which is either an increase from A) 1 to 1.25, B) 1.25 to 1.5, or C) 1.5 to 1.75. The grey dotted line represents the threshold of $R_t = 1$. The estimates (plotted in the middle of each time window) and their 95% credible intervals (shaded area) are very similar and therefore overlap. D-F) The bias (or absolute difference) in the mean $R_t$ estimated from weekly data compared to the true value of $R_t$. The shaded area corresponds to the 95% quantiles of the bias across the 100 simulations. G-I) The uncertainty (mean width of the 95% credible interval) in the $R_t$ values estimated from weekly data. J-L) The proportion of the 100 $R_t$ estimates made using weekly data where the 95% credible interval encompasses either the true value of $R_t$ (black) or the value of $R_t$ that would have been estimated from daily data (blue).

e. **Influence of $R_t$ plotting time relative to the time window used for estimation**

By default, the EpiEstim R package estimates $R_t$ over weekly sliding time windows ending at time t, at which point $R_t$ is plotted. An alternative to this, is to plot $R_t$ in the middle of the time window, centred around t. This would mean that $R_t$ estimates cannot be made in real-time, i.e., if a weekly window was used you can only estimate $R_t$ at t + 3.5. However, the advantage is that retrospective $R_t$ estimates would be less influenced by the lag corresponding to the length of the time window.

To demonstrate this, we reconsider the scenarios with a gradual and sudden change in $R_t$. When gradual changes in $R_t$ are plotted in the middle of the time window (t-3.5), the true value of $R_t$ is well recovered and the 95% coverage is considerably improved in comparison to estimates plotted at the end of the time window (Figure S15). Overall, the 95% credible intervals of the gradual $R_t$ estimates plotted at the mid-point of the time window encompassed the true value of $R_t$ 93-97% and 88-96% of the time for the increasing and decreasing scenarios respectively (compared to 78-96% and 62-94% when $R_t$ is plotted at the end of the time window). In the gradual change from 1.5 to 1.75 and 1.75 to 1.5 scenarios, there is still a dip in coverage which occurs when case numbers are so high that the 95% credible intervals become extremely narrow (Figure S15C & J). Therefore, when the sliding window encompasses incidence data before $R_t$ plateaus, it means $R_t$ is slightly under- or overestimated for increasing and decreasing $R_t$ respectively. However, it is evident that the mean estimates are negligibly different to the true value, despite not being exactly the same.

In the stepwise change in $R_t$ scenarios, there is also some improvement in 95% coverage (Figure S16). There is still a lag corresponding to the length of the time window, but now the change in $R_t$ is detected earlier and the dip in coverage is centred around the time change on day 35. As above, the dips in coverage are larger when case numbers are very high, and the 95% credible intervals are small. The 95% credible interval of estimates plotted at the mid-point of the time window encompassed the true value of $R_t$ 88-95% and 85-93% of the time for stepwise increasing and decreasing scenarios respectively, which is a modest improvement compared to 87-95% and 85-90% when $R_t$ is plotted at the end of the time window.

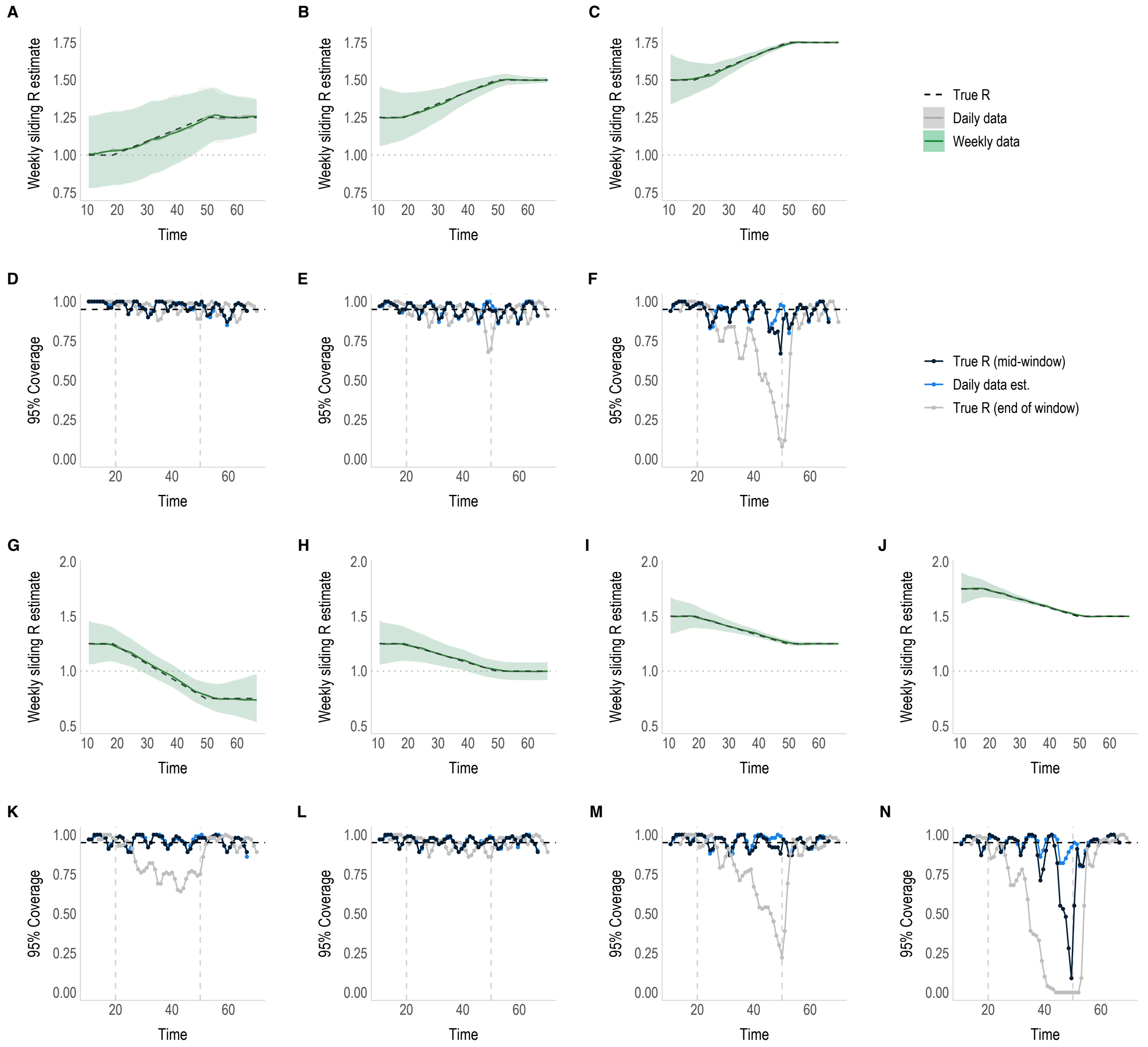

**Figure S15.** Plotting the gradual change in $R_t$ scenarios for 100 simulated epidemics in the middle of the time window. Here, the gradually increasing (A-C) and decreasing (G-J) $R_t$ estimate plots are exactly as in figures S13 and S14, except they are plotted at t-3.5. Each 95% coverage plot (D-F & K-N) corresponds to the plot directly above and shows the proportion of the 100 $R_t$ estimates made using weekly data where the 95% credible interval encompasses either the true value of $R_t$ (black) or the value of $R_t$ that would have been estimated from daily data (blue). As a reference, the 95% coverage of the true value of $R_t$ for the $R_t$ estimates plotted at the end of the time window (see figures S13 & S14) are shown in grey.

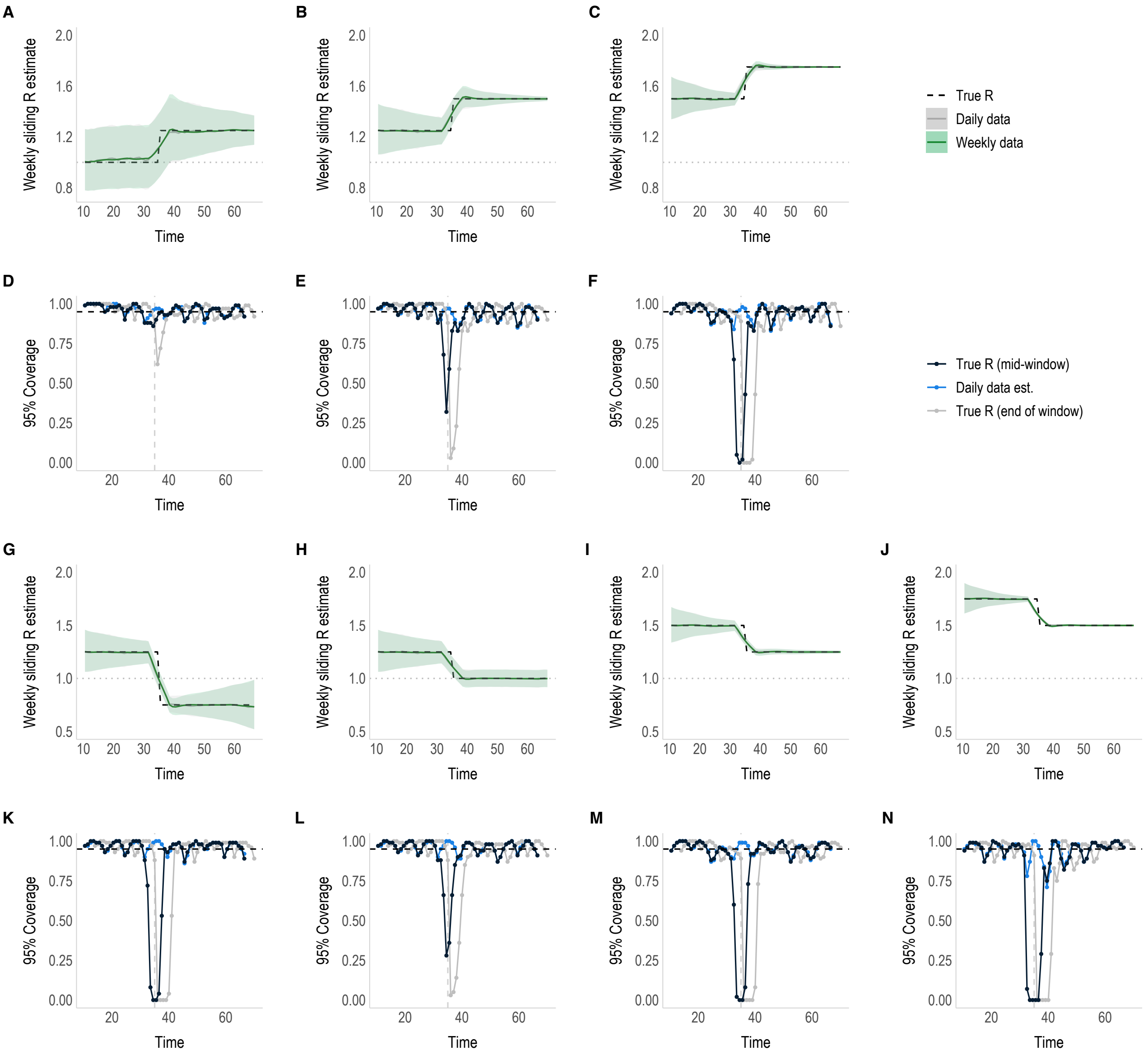

**Figure S16.** Plotting the stepwise change in $R_t$ scenarios for 100 simulated epidemics in the middle of the time window. Here, the stepwise increasing (A-C) and decreasing (G-J) $R_t$ estimate plots are exactly as in figures S11 and S12, except they are plotted at t-3.5. Each 95% coverage plot (D-F & K-N) corresponds to the plot directly above and shows the proportion of the 100 $R_t$ estimates made using weekly data where the 95% credible interval encompasses either the true value of $R_t$ (black) or the value of $R_t$ that would have been estimated from daily data (blue). As a reference, the 95% coverage of the true value of $R_t$ for the $R_t$ estimates plotted at the end of the time window (see figures S11 & S12) are shown in grey.

#### f. Weekend effects

To mimic incidence data with weekend effects, the simulated incidence for scenarios where $R_t$ remained constant at 1.5, suddenly decreased from 1.5 to 1.25, and gradually increased from 1.25 to 1.5, were further modified so that 80% of cases were taken off the final two days of each aggregation window and redistributed uniformly over the first two days of the aggregation window (Figure S17A-C). This appears roughly similar to the pattern observed in the influenza case study (Figure S7). $R_t$ was then estimated using daily (Figure S17D-F), weekly (Figure S17G-I), and two-weekly (Figure S17J-L) sliding windows, using the simulated data (with weekend effects) and the reconstructed daily data.

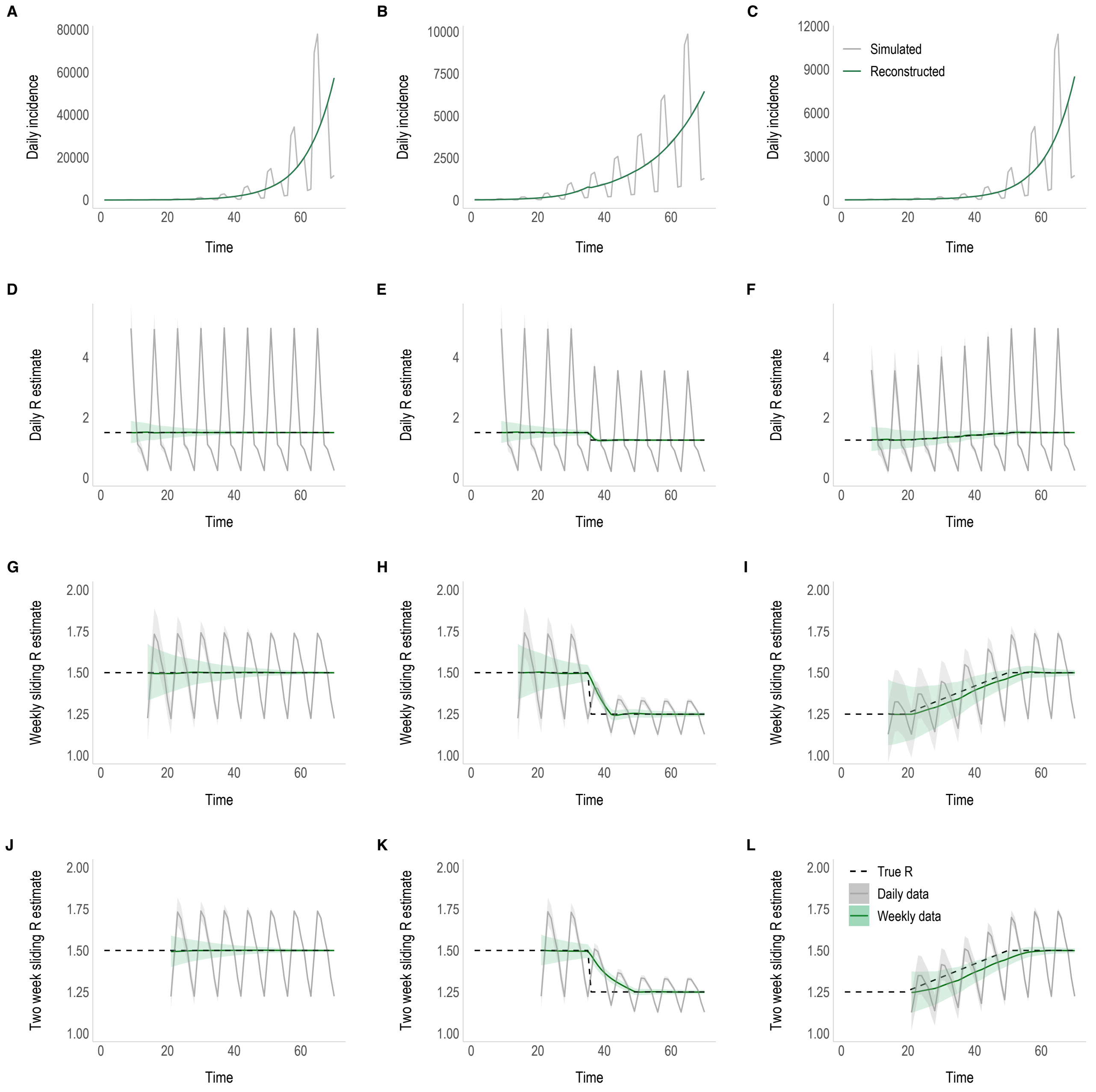

**Figure S17.** Assessing the performance of our method in the presence of weekend effects in the reported data. A-C) Example of reported (grey) and reconstructed (green) daily incidence data taken from one of the 100 simulated epidemics (selected at random) for a (A) constant $R_t$ of 1.5, (B) stepwise decrease in $R_t$ from 1.5 to 1.25, and (C) gradual increase in $R_t$ from 1.25 to 1.5. $R_t$ was estimated over D-F) daily, G-I) weekly, and J-L) two-weekly sliding time windows. The black dashed line is the true value of $R_t$ that the simulated data was based on. Note: y-axis limits for each incidence plot varies and the y-axis scale is different for the daily $R_t$ estimate plots (second row).

For every scenario, the $R_t$ estimates were considerably smoother and more accurate when the incidence had been reconstructed from weekly data. This is simply because the reconstruction smoothed out the intra-weekly variability and removed the effect of the noise from $R_t$ estimates.

#### g. Number of iterations

For each scenario, the EM algorithm used to reconstruct the incidence (and in turn estimate $R_t$) converged over a small number of iterations, with negligible differences beyond 5 iterations (Figure S18). Given that the computational time is so short (see section 2f), the default number of iterations was set to 10 in the R package, although the user can change that setting.

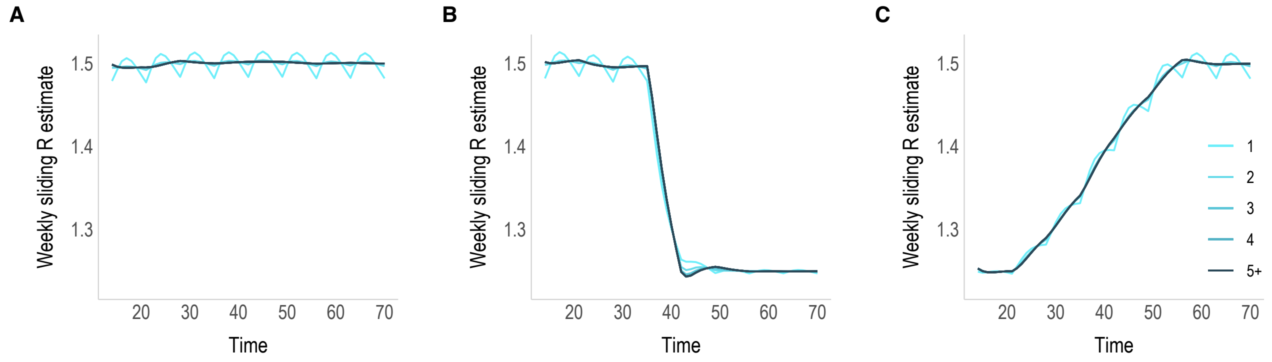

**Figure S18.** The mean weekly sliding $R_t$ estimates generated after each of the 10 iterations from 100 stochastically simulated epidemics for the following scenarios: A) constant $R_t$ of 1.5, B) stepwise decrease in $R_t$ from 1.5 to 1.25, and C) gradual increase in $R_t$ from 1.25 to 1.5.

#### h. Different temporal aggregations

In addition to weekly incidence, the method can be successfully applied to other temporal aggregations of data. Here, we showcase the method applied to data that has been aggregated to 3-day, 10-day and 14-day timescales. First, we consider the ideal scenario where the end of the aggregation window aligns perfectly with the $R_t$ step-change on day 35 (Figure S19). Then we consider how the accuracy of the estimations would be affected if the step-change fell in the middle of the aggregation window (Figure S20).

When the aggregation window is aligned with the step-change, there is only the usual lag in detecting temporal changes in $R_t$, corresponding to the width of the sliding time window used for the estimation (Figure S19 B, E, H & K). However, when aggregation windows misalign with the step-change, there is a further lag (Figure S20 H & K). This is because, in the process of daily incidence reconstruction, the growth rate is assumed constant within each aggregation window. When aggregation windows misalign with the step-change, the growth rate is essentially smoothed out when the daily incidence is reconstructed. This is important to be aware of when using larger aggregations of data, as the loss of some temporal resolution in $R_t$ estimates will be unavoidable.

As mentioned in section 2b, we recommend that the user ensures that the sliding window used to estimate $R_t$ is equal to or longer than the length of the aggregation window. Although a longer sliding window leads to a further lag in $R_t$ estimates for the stepwise change in $R_t$ scenarios (Panels H & K in Figures S19 & S20), the $R_t$ estimates generated are smoother. This is because they are less affected by discontinuities in the reconstructed incidence data, the result of which can be clearly seen in the gradual change in $R_t$ scenarios (Panels I & L in Figures S19 & S20), where estimates made using 7-day sliding windows appear wavy.

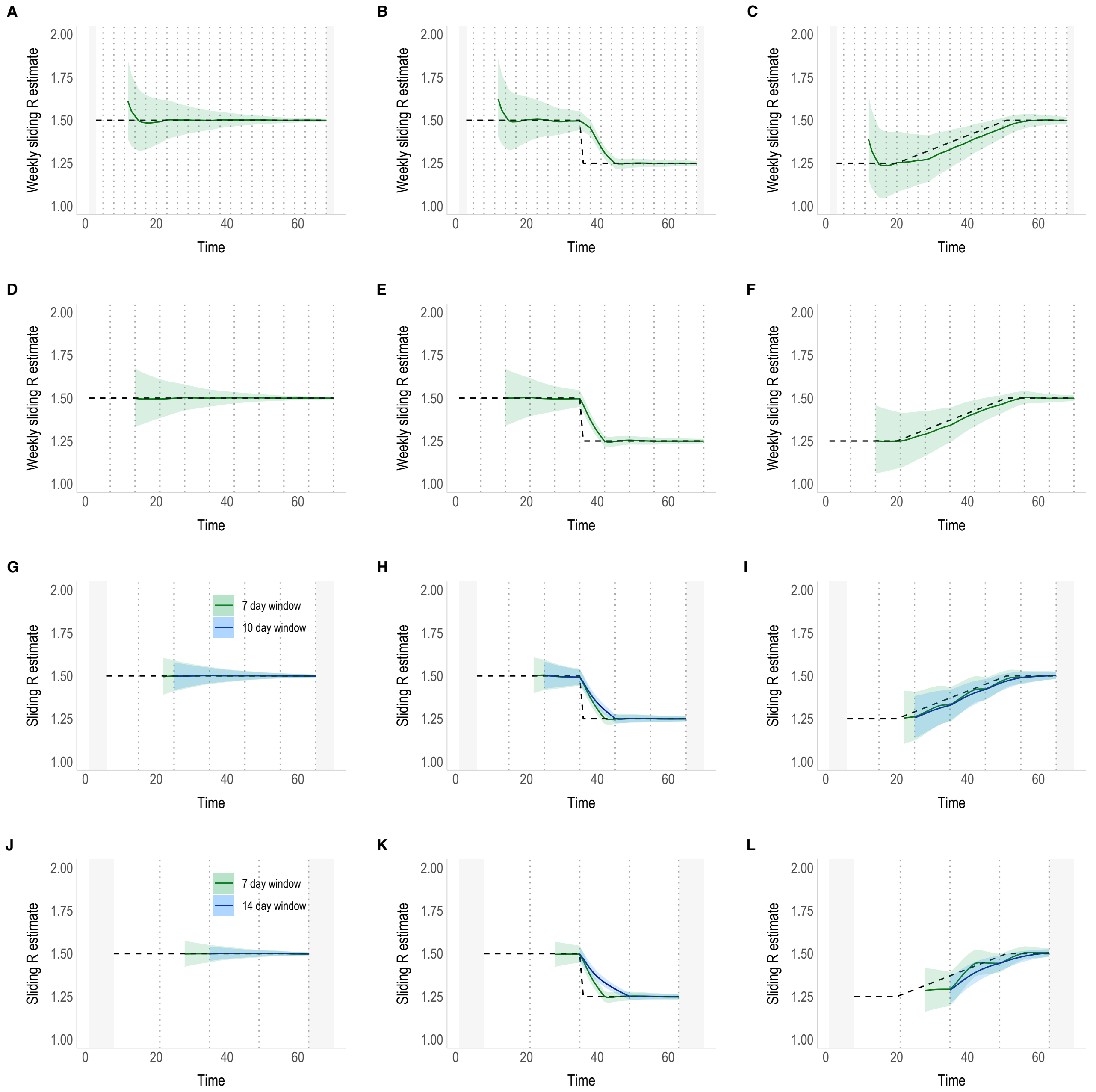

**Figure S19.** Performance of the method using alternative aggregations of incidence data when the end of aggregation windows align with the step change on day 35. We consider the following scenarios: a constant $R_t$ of 1.5 (left), stepwise decrease in $R_t$ from 1.5 to 1.25 (middle), and a gradual increase in $R_t$ from 1.25 to 1.5 (right), with data aggregated over A-C) 3 days, D-F) 7 days, G-I) 10 days, and J-L) 14 days. For the 10-day and 14-day aggregations of data, we compare estimates made using weekly sliding windows (green) with sliding windows of length matching that of the aggregation of data (blue). For all scenarios, $R_t$ estimates are plotted at the end of the sliding time window. Grey dotted lines show the end of each aggregation window and the grey shaded areas are the time periods excluded in order to align the aggregation windows (if necessary).

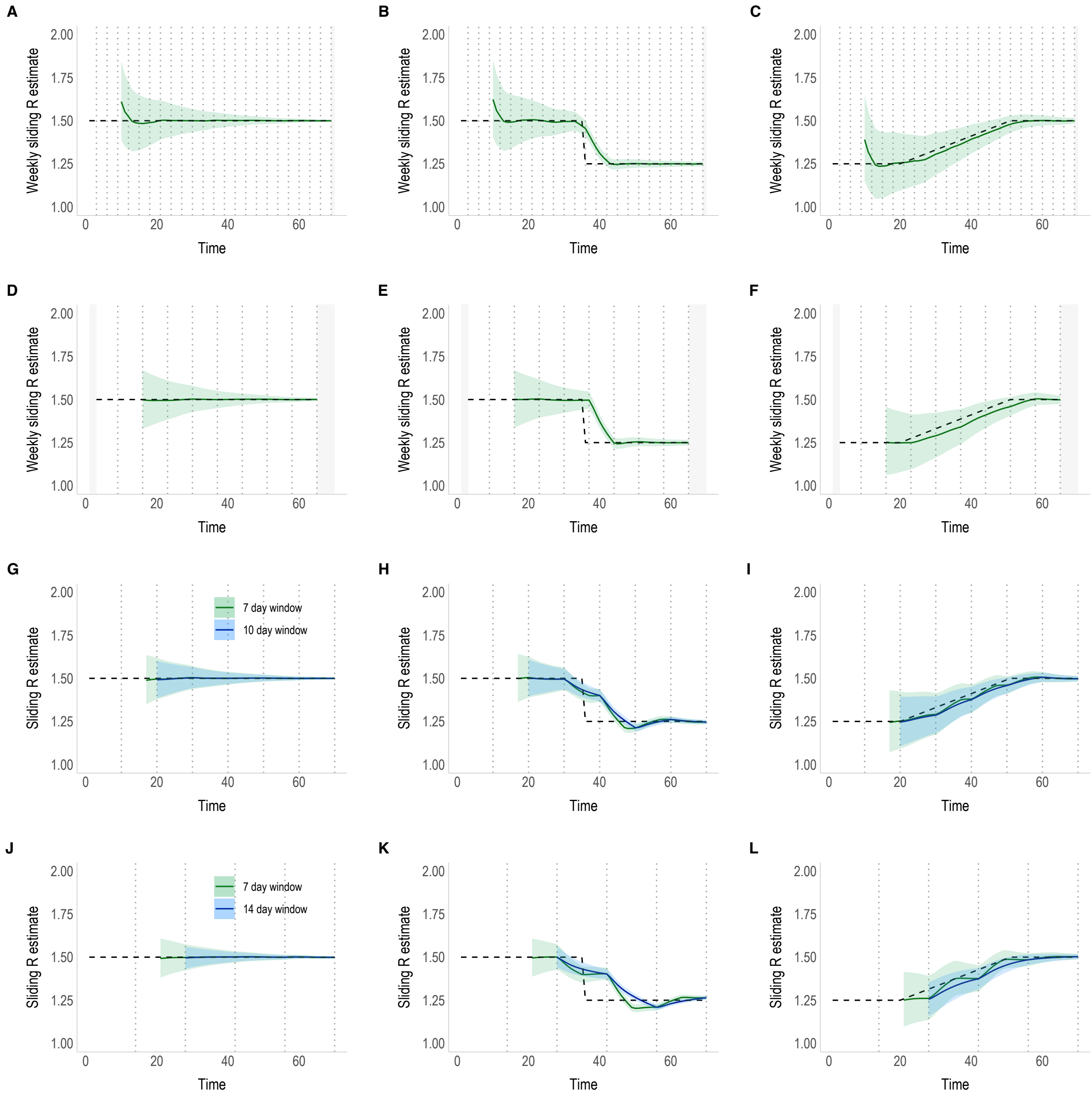

**Figure S20.** Performance of the method using alternative aggregations of incidence data when the end of aggregation windows <u>do not align</u> with the step change on day 35. We consider the following scenarios: a constant $R_t$ of 1.5 (left), stepwise decrease in $R_t$ from 1.5 to 1.25 (middle), and a gradual increase in $R_t$ from 1.25 to 1.5 (right), with data aggregated over A-C) 3-days, D-F) 7-days, G-I) 10-days, and J-L) 14-days. For the 10-day and 14-day aggregations of data, we compare estimates made using weekly sliding windows (green) with sliding windows of length matching that of the aggregation of data (blue). For all scenarios, $R_t$ estimates are plotted at the end of the sliding time window. Grey dotted lines show the end of each aggregation window and the grey shaded areas are the time periods excluded in order to misalign the aggregation windows (if necessary).

### References

1. Cori A, Ferguson NM, Fraser C, Cauchemez S. A New Framework and Software to Estimate Time-Varying Reproduction Numbers During Epidemics. Am J Epidemiol. 2013 Nov 1;178(9):1505–12.

2. Riley P, Cost AA, Riley S. Intra-Weekly Variations of Influenza-Like Illness in Military Populations. Mil Med. 2016 Apr 1;181(4):364–8.

3. Jombart T, Nouvellet P, Bhatia S, Kamvar ZN, Taylor T, Ghozzi S. projections: Project Future Case Incidence [Internet]. 2021 [cited 2022 Jun 9]. Available from: https://CRAN.R-project.org/package=projections
